# Age norms for DunedinPACE: An epigenetic pace of aging biomarker

**DOI:** 10.64898/2026.08.13.26360306

**Authors:** Kyle J. Bourassa, Calen P. Ryan, Karen Sugden, Ethan T. Whitman, Melanie E. Garrett, Renate M. Houts, Claire E. Indik, William Marella, Benjamin S. Williams, VA Mid Atlantic MIRECC Workgroup, Allison E. Aiello, Kathleen Mullan Harris, David L. Corcoran, Allison E. Ashley-Koch, Jean C. Beckham, Nathan A. Kimbrel, Ahmad R. Hariri, Avshalom Caspi, Terrie E. Moffitt, Daniel W. Belsky

## Abstract

Epigenetic clocks have transformed the study of biological aging in epidemiology and clinical trials. However, the utility of these measures in clinical settings is limited by a lack of population-based norms that clinicians, patients, and researchers can use to understand and communicate how fast an individual is aging relative to same-aged peers. Here, we developed age norms for DunedinPACE, an epigenetic Pace of Aging measure derived from DNA methylation. To do so, we meta-analyzed data from 11 cohorts (*N* = 37,855 individuals, ages 17-99 years) to characterize the association between chronological age and DunedinPACE. We investigated sex differences and nonlinearity, confirmed results using longitudinal data, verified that age-normed DunedinPACE scores predict clinical outcomes, and illustrated how norms support the needs of clinical aging research. The age norms reported here will help integrate biomarkers of aging, such as DunedinPACE, into precision public health and medicine.

## Introduction

Epigenetic clocks are biomarkers of aging^1–4^. They aim to measure the pace and progress of biological changes that undermine the integrity and resilience of our cells, tissues, and organs as we grow older^5–7^. The best-validated epigenetic clocks predict healthspan and lifespan in populations around the world^8^. On the strength of this evidence, epigenetic clocks are now used to study how environments and behaviors affect aging processes^9–10^. In parallel, a growing number of clinical trials are using epigenetic clocks to evaluate how behavioral, lifestyle, and pharmacological interventions alter aging biology to increase healthspan^11–13^. However, applications of aging biomarkers in clinical research and their adoption in clinical care remain limited. One barrier to the clinical translation of epigenetic clocks is the challenge of interpreting individual scores. Clinicians interpret results for normative measures using clinical reference ranges or population-based growth standards; e.g., pediatricians use percentile ranks to interpret children’s heights and weights and communicate their meaning to parents. Without the ability to compare to a reference population, it is difficult to know whether a patient’s aging score signifies healthy or unhealthy aging. This challenge is particularly relevant to epigenetic clocks that measure the Pace of Aging, where a rate of aging is difficult to interpret without reference norms.

Most epigenetic clocks estimate a person’s biological age, which represents their biological risk for aging-related mortality disease, disability, and mortality^14–17^. These clocks function like odometers for the aging process, indexing the extent of aging-related biological decline a person has accumulated from conception to the point of sample collection. In this cumulative approach, values older than a person’s calendar age indicate advanced biological aging, whereas values younger than a person’s calendar age indicate delayed aging. In contrast to odometer-type clocks, Pace of Aging epigenetic clocks function as speedometers for the aging process, quantifying a person’s current rate of aging-related biological deterioration^18–19^. This rate of change value represents a ratio of years of biological change experienced for each 12 months of calendar time. Values below one indicate a slower Pace of Aging relative to a midlife reference norm; values above one indicate a faster Pace of Aging.

Pace of Aging clocks stand out in aging research for their sensitivity to adverse environmental exposures, ability to predict clinical outcomes, and responsiveness to intervention^11–12,20–24^. However, the values present challenges for clinical interpretation. DunedinPACE is built around a reference value of 1.00, aligned to the average Pace of Aging observed among 45-year-old members of the Dunedin Study birth cohort. The challenge arises because the rate at which people age biologically tends to accelerate as they grow older. This acceleration in the rate of aging is observed in demography, where risk of death doubles roughly every eight years from the fourth decade of life onward^25^. In medicine, declines in organ system integrity, physical function, and cognitive performance, as well as risk for incident chronic disease, appear to accelerate across midlife through the end of the life course^26–27^. In geroscience, new methods are revealing that shifts in molecular profiles accelerate as patients grow older^28–30^. Consistent with these patterns, Pace of Aging, indexed by DunedinPACE, tends to be faster in older people when compared to younger people^19^. This means the same Pace of Aging score could indicate faster-than-normal aging for a younger person and slower-than-normal aging for an older person.

To facilitate translation of Pace of Aging epigenetic clocks into clinical settings, reference norms are needed to enable straightforward interpretation of scores for patients of any age. Here, we establish age norms for DunedinPACE using data from 37,855 individuals across 11 cohorts with diverse genetic ancestry, nationality, and geography. Across these cohorts, we evaluated potential sex differences and nonlinearity, validated the results against longitudinal within-person change, and tested sensitivity to cell composition of the DNA samples and to different generations of DNA methylation microarrays. We also confirmed that age-normed DunedinPACE values retain predictive power for clinical outcomes and illustrate applications for this approach. Finally, we provide a web-based interface to calculate percentile ranks and code to normalize DunedinPACE aging scores.

## Methods

### Participants and Study Design

We analyzed data from 11 cohorts that included information from 37,855 individuals across 43,010 observations. The cohorts included: the Health and Retirement Study (HRS), Coronary Artery Risk Development in Young Adults Study (CARDIA), Women’s Health Initiative (WHI), Framingham Heart Study 3^rd^ Generation (FHS 3^rd^ Gen) and FHS Offspring studies, Swedish Adoption/Twin Study of Aging (SATSA), Detroit Neighborhood Health Study (DNHS), National Longitudinal Study of Adolescent to Adult Health (Add Health), Midlife Development in the US Study (MIDUS), Post-Deployment Mental Health Study (PDMH), and Generation Scotland. Details for cohorts are presented in Supplemental Table 1.

### Measures

#### DNA methylation (DNAm) data

DNAm data were derived from blood using the Illumina 450k and EPIC v1.0 arrays (Illumina Inc., San Diego, CA). Established methods were used to estimate white blood cell composition^31–33^. Details for DNA methylation processing and normalization in the cohorts^34^ are reported in Supplemental Table 2.

### Epigenetic Pace of Aging

We assessed Pace of Aging using DunedinPACE scores calculated from DNAm data using the DunedinPACE R package^35^.

### Data Analysis

We first quantified associations between chronological age and DunedinPACE in regression models. We used generalized estimating equations to account for the non-independence of repeated measures in three cohorts with longitudinal DNAm data (CARDIA, DNHS, SATSA). Results were modeled within each cohort and integrated using random-effects meta-analysis. Additional details are provided in Supplemental Methods 1. Second, we tested sex differences by comparing estimates of associations between men and women using subgroup analysis. Third, we tested nonlinearity by meta-analyzing the age association for higher-order terms (e.g., age^2^, age^3^), as well as by investigating changes in variance explained and model fit when including higher-order terms. Fourth, we replicated the parameters estimated in the between-person comparisons using fixed-effects (within-person) regression models in cohorts with longitudinal DNAm data. Finally, we examined differences in associations based on the generation of microarray used to measure DNAm data and tested associations after correcting for the cell composition of the DNA samples.

## Results

The 37,855 participants (57.8% women) averaged 50.9 years old (SD = 10.7, 17-99 years). DunedinPACE values averaged 1.00 biological year of decline per chronological year (SD = 0.13; range 0.37 to 1.75; Table 1; Figure 1).

**Figure 1.**
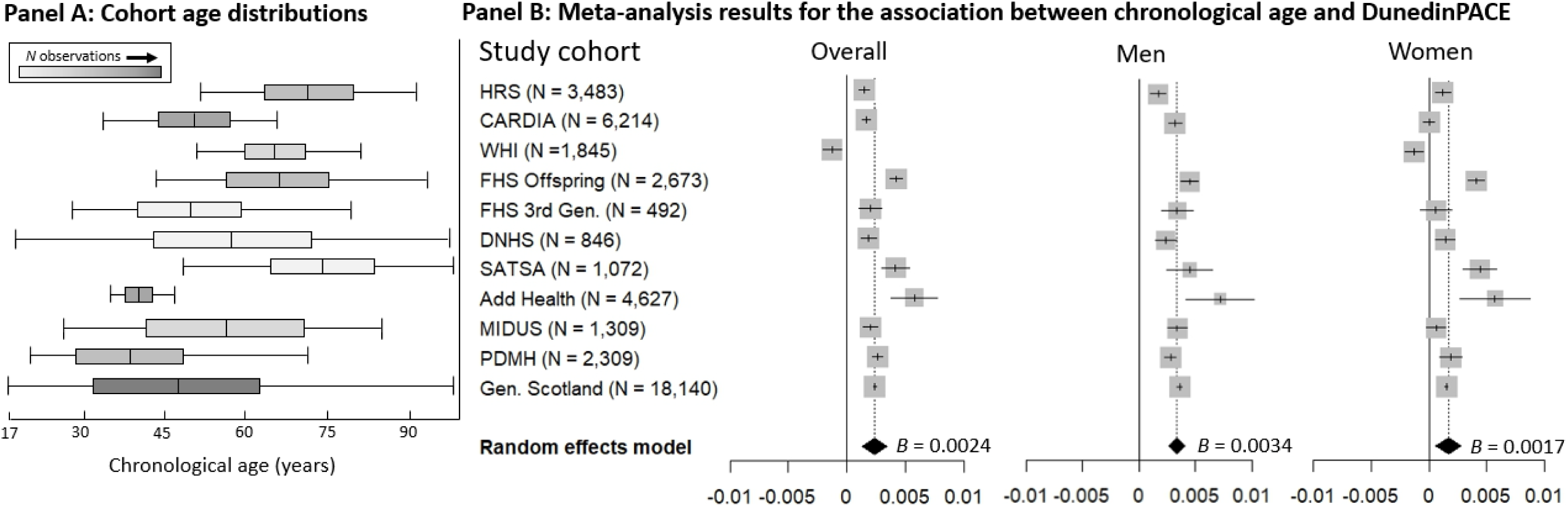
Visualization of cohorts’ age distributions and forest plots of the association between chronological age and DunedinPACE. **Panel A** shows a conceptual illustration of the age distributions for the different cohorts. Boxes represent means and 1 *SD* of age above and below the mean, whiskers represent the range of ages, and darker shading represents relatively more observations. **Panel B** shows the meta-analyzed results of the association between chronological age and DunedinPACE for the full cohorts, as well as for men and women only. *N*s describe total observations by cohort, *n*s for men and women are provided in Table 2. Grey squares represent the contribution of each study to the overall estimate, shown by the black diamond. Whiskers represent 95% confidence intervals.

**Table 1.** Cohort characteristics.

| Cohort | Obs. | n | % male | Chronological Age |  | DunedinPACE |  |
| --- | --- | --- | --- | --- | --- | --- | --- |
|  |  |  |  | Mean (SD) | Range | Mean (SD) | Range |
| HRS<sup>A</sup> | 3,483 | 3,483 | 41.2% | 69.6 (9.5) | 50-91 | 1.028 (0.14) | 0.64-1.54 |
| CARDIA<sup>A</sup> | 6,214 | 2,023 | 42.6% | 48.8 (6.5) | 32-64 | 0.976 (0.13) | 0.37-1.56 |
| WHI<sup>B</sup> | 1,845 | 1,845 | 0.0% | 64.2 (7.0) | 50-79 | 1.020 (0.12) | 0.58-1.41 |
| FHS Offspring<sup>A</sup> | 2,673 | 2,673 | 45.6% | 66.4 (8.9) | 40-92 | 0.972 (0.13) | 0.46-1.60 |
| FHS 3rd Gen.<sup>A</sup> | 492 | 492 | 46.7% | 48.2 (9.7) | 25-78 | 0.928 (0.11) | 0.65-1.32 |
| DNHS<sup>A</sup> | 846 | 510 | 40.6% | 56.3 (15.9) | 18-98 | 1.113 (0.13) | 0.77-1.59 |
| SATSA<sup>B</sup> | 1,072 | 444 | 40.5% | 73.1 (9.8) | 48-99 | 1.062 (0.17) | 0.61-1.75 |
| Add Health<sup>A</sup> | 4,627 | 4,627 | 39.8% | 38.5 (1.9) | 33-45 | 0.968 (0.13) | 0.37-1.59 |
| MIDUS<sup>A</sup> | 1,309 | 1,309 | 44.2% | 54.0 (12.6) | 26-86 | 0.990 (0.14) | 0.53-1.45 |
| PDMH<sup>A,B</sup> | 2,309 | 2,309 | 78.7% | 37.4 (10.4) | 20-70 | 1.067 (0.11) | 0.75-1.57 |
| Generation Scotland<sup>A</sup> | 18,140 | 18,140 | 41.2% | 47.6 (14.9) | 17-99 | 0.980 (0.13) | 0.41-1.62 |
| **Totals** | **43,010** | **37,855** | **42.4%** | **50.9 (10.7)** | **17-99** | **1.001 (0.13)** | **0.37-1.75** |

*Note:* Observation and *N* totals = sums, % male, mean, and *SD* totals = weighted averages, range total = most extreme values. Obs. = observations, *SD* = Standard deviation. Values for sex, age, and DunedinPACE are weighted based on sample size for the cohorts. HRS = Health and Retirement Study, CARDIA = Coronary Artery Risk Development in Young Adults Study, WHI = Women's Health Initiative, FHS = Framingham Heart Study, DNHS = Detroit Neighborhood Health Study, SATSA = Swedish Adoption/Twin Study of Aging, Add Health = National Longitudinal Study of Adolescent to Adult Health, MIDUS = Midlife Development in the US, PDMH = Post-Deployment Mental Health Study.
<sup>A</sup> = EPIC v1.0 chip used in sample, <sup>B</sup> = 450K chip used in sample

**Table 2.** Association between chronological age and DunedinPACE.

| Cohort | Obs. | Overall association between age and DunedinPACE |  | Association among men |  | Association among women |  |
| --- | --- | --- | --- | --- | --- | --- | --- |
|  |  | *B* | 95% CI | *B* | 95% CI | *B* | 95% CI |
| HRS | 3,483 | 0.0015 | 0.0010, 0.0020 | 0.0017 | 0.0010, 0.0024 | 0.0013 | 0.0006, 0.0019 |
| CARDIA | 6,214 | 0.0017 | 0.0013, 0.0021 | 0.0032 | 0.0026, 0.0038 | 0.0007 | 0.0002, 0.0012 |
| WHI | 1,845 | -0.0013 | -0.0021, -0.0005 | — | — | -0.0013 | -0.0021, -0.0005 |
| FHS Offspring | 2,673 | 0.0042 | 0.0037, 0.0048 | 0.0045 | 0.0037, 0.0052 | 0.0041 | 0.0034, 0.0048 |
| FHS 3rd Gen. | 492 | 0.0020 | 0.0010, 0.0030 | 0.0034 | 0.0020, 0.0027 | 0.0006 | -0.0008, 0.0019 |
| DNHS | 846 | 0.0019 | 0.0012, 0.0025 | 0.0024 | 0.0014, 0.0034 | 0.0014 | 0.0005, 0.0022 |
| SATSA | 1,072 | 0.0042 | 0.0030, 0.0054 | 0.0045 | 0.0025, 0.0065 | 0.0044 | 0.0029, 0.0059 |
| Add Health | 4,627 | 0.0058 | 0.0038, 0.0078 | 0.0072 | 0.0042, 0.0102 | 0.0057 | 0.0031, 0.0083 |
| MIDUS | 1,309 | 0.0020 | 0.0014, 0.0026 | 0.0034 | 0.0026, 0.0042 | 0.0006 | -0.0002, 0.0014 |
| PDMH | 2,309 | 0.0026 | 0.0022, 0.0031 | 0.0028 | 0.0023, 0.0033 | 0.0019 | 0.0010, 0.0029 |
| Generation Scotland | 18,140 | 0.0024 | 0.0023, 0.0025 | 0.0036 | 0.0034, 0.0038 | 0.0015 | 0.0014, 0.0017 |
| **Meta-analyzed results** |  | 0.0024 | 0.0014, 0.0034 | 0.0034 | 0.0027, 0.0041 | 0.0017 | 0.0005, 0.0028 |

*Note:* Meta-analyzed results were generated using a random effects model. WHI did not include any men in the cohort. Obs. = observations.

### Pace of Aging is Faster for Older Adults Compared to Younger Adults

Older participants had faster DunedinPACE aging scores when compared to younger participants; each decade of calendar age was associated with a 2.4% increase in the Pace of Aging (B = 0.0024, 95% CI [0.0014-0.0034], p < .001; Table 2; Figure 1).

### Pace of Aging Increases Faster for Men Compared to Women

Men have shorter lifespans than women and experience earlier onset of many aging-related diseases, suggesting sex differences in the pace of biological aging^36^. DunedinPACE accounted for such potential differences by using sex-specific reference norms^19,37^. However, this approach does not rule out differences in how the rate of aging might change for men and women across the lifespan. To test this possibility, we compared associations between age and DunedinPACE for men and women. Men showed a more rapid increase in DunedinPACE across the adult life span as compared with women (for men, B = 0.0034, 95% CI [0.0027-0.0041], p < .001; for women, B = 0.0017, 95% CI [0.0005-0.0028], p = .004; test of subgroup differences, Q_between_(1) = 6.35, p= 0.012). On average, these estimates suggest that men experience an increase of 3.4% in the rate of aging for each decade of age, as compared to an increase of 1.7% per decade of age for women.

### Pace of Aging Increases Linearly Over the Adult Lifespan

A linear association between age and a rate of aging implies nonlinear (accelerating) deterioration over time. However, it is possible that changes in the rate of aging are nonlinear. We tested this possibility by adding higher-order terms (i.e. age^2^ and age^3^) to our models and meta-analyzing the coefficients. We also tested changes in model fit and variance explained by these higher-order terms, and evaluated all models stratified by sex. Higher-order terms were not statistically significant in meta-analyses (all ps > .18), nor did they improve model fit or explain additional variance (Supplemental Text 1, Supplemental Tables 3-5). These results suggest that a linear model is sufficient to explain lifespan variation in DunedinPACE.

### Longitudinal Repeated Measures Analysis Confirms Accelerating Pace of Aging Over the Adult Lifespan

Using cross-sectional data to characterize associations at different ages can bias results due to generational differences in exposures that affect health and the epigenome (i.e., cohort effects). In cross-sectional data, age differences are perfectly correlated with differences in birth year. This makes it possible that the faster DunedinPACE aging scores apparent in older compared with younger people reflect a difference in the aging rate between earlier and more recently born cohorts^38^. To evaluate whether cohort effects might contaminate our estimates of age patterning, we repeated our analysis with three cohorts that collected DNA methylation data from participants at repeated intervals (combined N = 2,977; 8,132 observations; ages 18 to 99 years old; Supplemental Table 6). Our estimate of within-individual change was not statistically different from our estimate of between-individual differences (within-person: B = 0.0039, 95% CI [0.0015-0.0064], p < .001; between-person: B = 0.0024, 95% CI [0.0014-0.0034], test of subgroup differences, Q_between_(1) = 1.32, p = .25). These results provide evidence that acceleration of DunedinPACE scores across the adult lifespan is not an artifact of generational cohort effects.

### Accounting for Generations of Methylation Arrays

We tested the potential for technical artifacts arising from different generations of microarrays used to generate DNAm data in our analysis, specifically the Illumina Infinium Bead Chip 450k and EPIC v1.0 arrays. DunedinPACE associations with age were similar across arrays (450K B = 0.0018, 95% CI [-0.0013-0.0050]; EPIC v1.0 B = 0.0025, 95% CI [0.0018-0.0033]; test of subgroup differences, Q_between_(1) = 0.18, p = .67). This difference does not distinguish artifacts arising from array chemistries from differences in characteristics of the cohorts in which they were used. A more focused comparison would contrast associations estimated from 450K and EPIC v1.0 chips in the same study. In the PDMH cohort, which generated data using both generations of arrays, the association between age and DunedinPACE was consistent across arrays (450K B = 0.0027; EPIC v1 B = 0.0027).

### Accounting for Cell Compositions of DNA samples

Different cell types have different patterns of DNA methylation. The cellular composition of blood varies between individuals as a function of aging, representing a potential confound in DNA methylation analyses^39^. We tested if differences in the cellular composition of blood DNA samples might account for DunedinPACE associations with age. Adjusting for estimated cell type proportions partially attenuated the association relative to our original estimates, but it remained statistically different from zero (B = 0.0015 [0.0005-0.0025], p = 0.003; Supplemental Table 7). There is a lack of consensus about whether cell composition of blood is a component of the aging phenotype or a confound^31^ and our goal is to produce norms that can be applied at the individual level without covarying for measures like cell composition. Therefore, we rely on unadjusted analyses to formalize age norms.

### Generating Age-Normed Reference Scores to Aid Interpretation of Individual Aging Scores

We used sex-specific linear age parameters to calculate age-normed reference scores for DunedinPACE from ages 20 to 90 years (Figure 2). Age-normed reference scores represent the expected Pace of Aging across these ages (Supplemental Table 8, Supplemental Text 1). We combined age-normed reference scores with the variance parameter estimated from the data (a standard deviation of 0.13 that was consistent across cohorts, sex, and age; Supplemental Table 9) to calculate z-scores (Supplemental Text 2). These z-scores can be converted to percentile ranks that provide a direct comparison to same-aged peers. Confidence intervals for percentile ranks were calculated based on a standard error of measurement for DunedinPACE^19^ (Supplemental Text 2).

**Figure 2.**
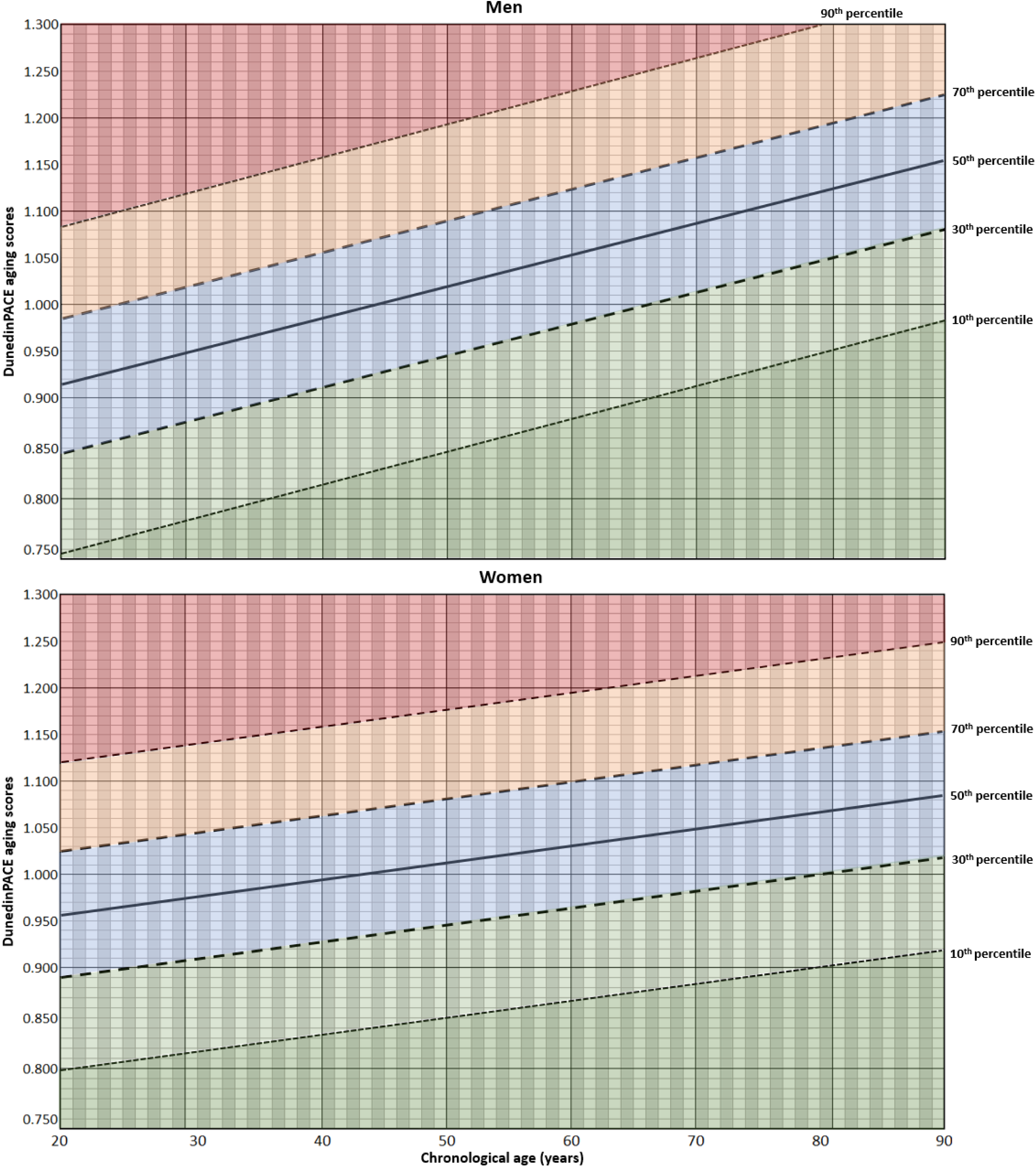
Age-normed DunedinPACE scores for men and women across the adult lifespan. Sex-specific linear age parameters and original DunedinPACE reference score for 45-year-olds (1.00) produced age-normed reference scores (see **Supplemental Text 2**) used to generate pace of aging z-scores and percentile ranks from age 20 to 90 years (see **Supplemental Table 8**). Overall normed values are shown in **Supplemental Figure 1**.

We also created a simple calculation that can normalize DunedinPACE scores to a reference value of 1.00 regardless of an individual’s age to allow for more direct comparison of scores between individuals of different ages:

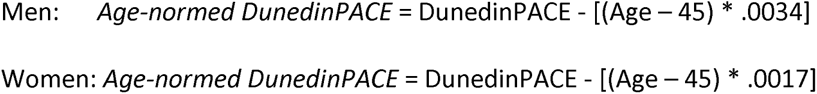

These normed scores are useful in situations where controlling for age statistically is not possible, such as in some clinical research settings.

Calculations can be made using our web application to generate percentile ranks and 95% confidence intervals for DunedinPACE values (https://tinyurl.com/DunedinPACE-Norms) or within the updated DunedinPACE R package, available on GitHub^28^ (https://github.com/danbelsky/DunedinPACE).

### Predicting Clinical Outcomes Using Age-Normed DunedinPACE

We used clinical outcome data from two cohorts (PDMH, MIDUS) to test whether age-normed DunedinPACE scores were associated with future chronic disease and death. As illustrated in Figure 3, age-normed DunedinPACE scores predicted clinical outcomes (e.g., chronic disease onset, all-cause mortality) with effect sizes comparable to those observed for the original, unadjusted DunedinPACE scores (Supplemental Table 10).

**Figure 3.**
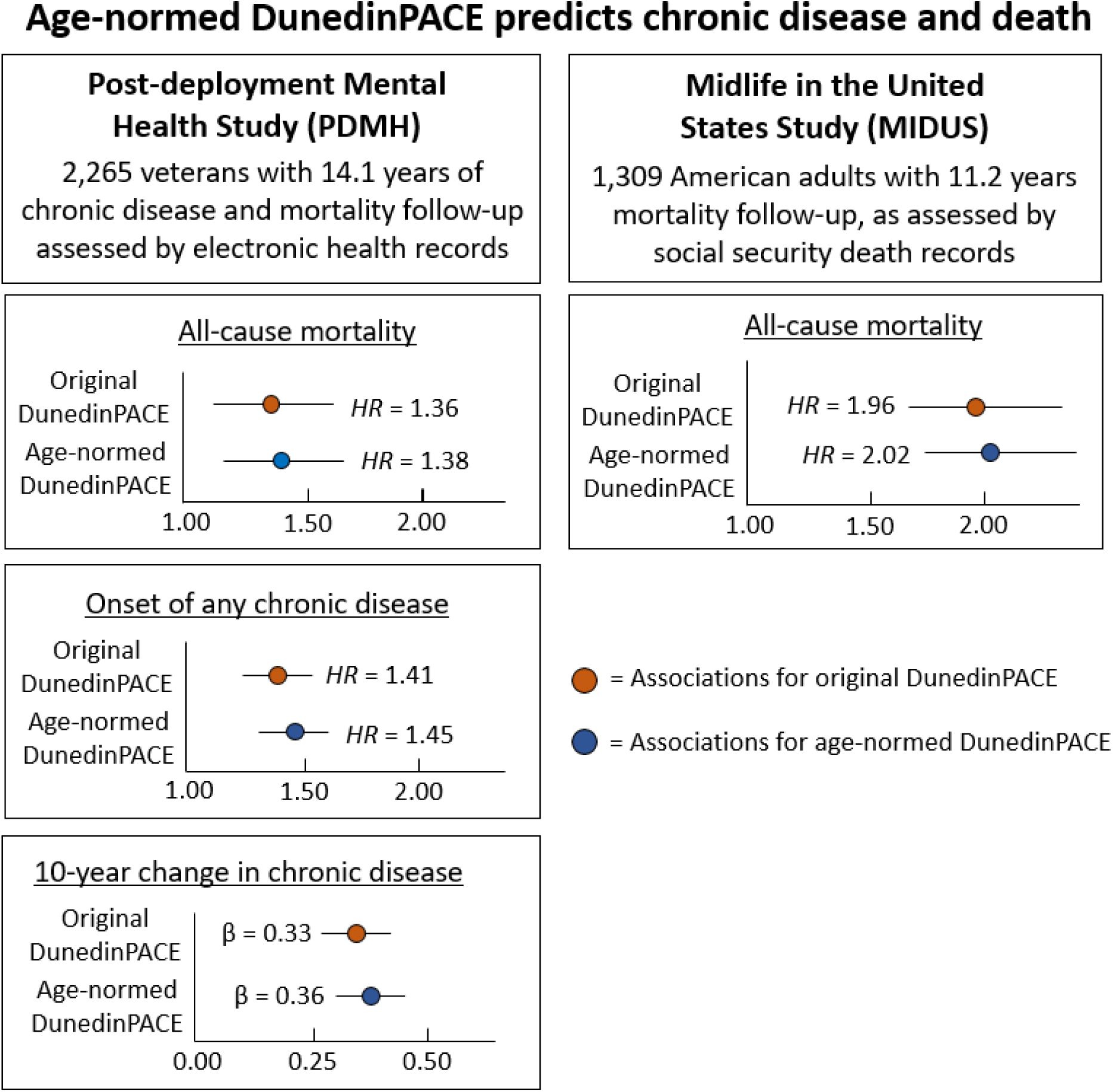
Age-normed DunedinPACE predicts chronic disease and death. This figure compares associations between original and age-normed DunedinPACE scores and clinical outcomes. All model estimates were adjusted for chronological age. Whiskers represent 95% confidence intervals and. In the PDMH, we used an average of 14.1 years of electronic health records to ascertain three clinical outcomes: (a) all-cause mortality, (b) chronic disease onset, and (c) change in chronic disease burden assessed by Charlson Comorbidity Index scores. In the MIDUS study, we assessed mortality ascertained to the end of 2023, n average follow-up of 11.2 years (**Supplemental Methods 2** provides full details for both cohorts). The effect sizes were largely comparable (**Supplemental Table 10** provides full estimates).

### Aiding Clinical Interpretation Using Age-Normed DunedinPACE

Normed values provide the opportunity to compare any individual’s DunedinPACE scores to the expected scores for same-age and same-sex peers. For example, a 25-year-old man with a DunedinPACE of 0.988 might at first appear to be aging more slowly than the original DunedinPACE reference score (e.g., 1.00). However, our age-norming calculation shows an expected aging score of 0.932 for a 25-year-old man, indicating he should be interpreted as aging 0.43 SD more quickly than expected for a man his age, placing him in the 66^th^ percentile. Similarly, an 80-year-old woman with a DunedinPACE of 1.05 might at first appear to be aging more quickly than the original DunedinPACE reference score. However, she would be interpreted as aging 0.08 SD more slowly than expected given the normed DunedinPACE score for women her age (1.060), placing her in the 47^th^ percentile.

Normed values can also assist with interpreting clinical research findings in unique populations that might not have an unexposed control group to provide a direct comparison. Figure 4 presents two illustrations of how age-norming DunedinPACE scores can aid in the interpretation of research findings, specifically drawing on published studies of a clinical sample of individuals with sickle cell disease and a cohort of older adults who were almost 90 years old on average^22,40^.

**Figure 4.**
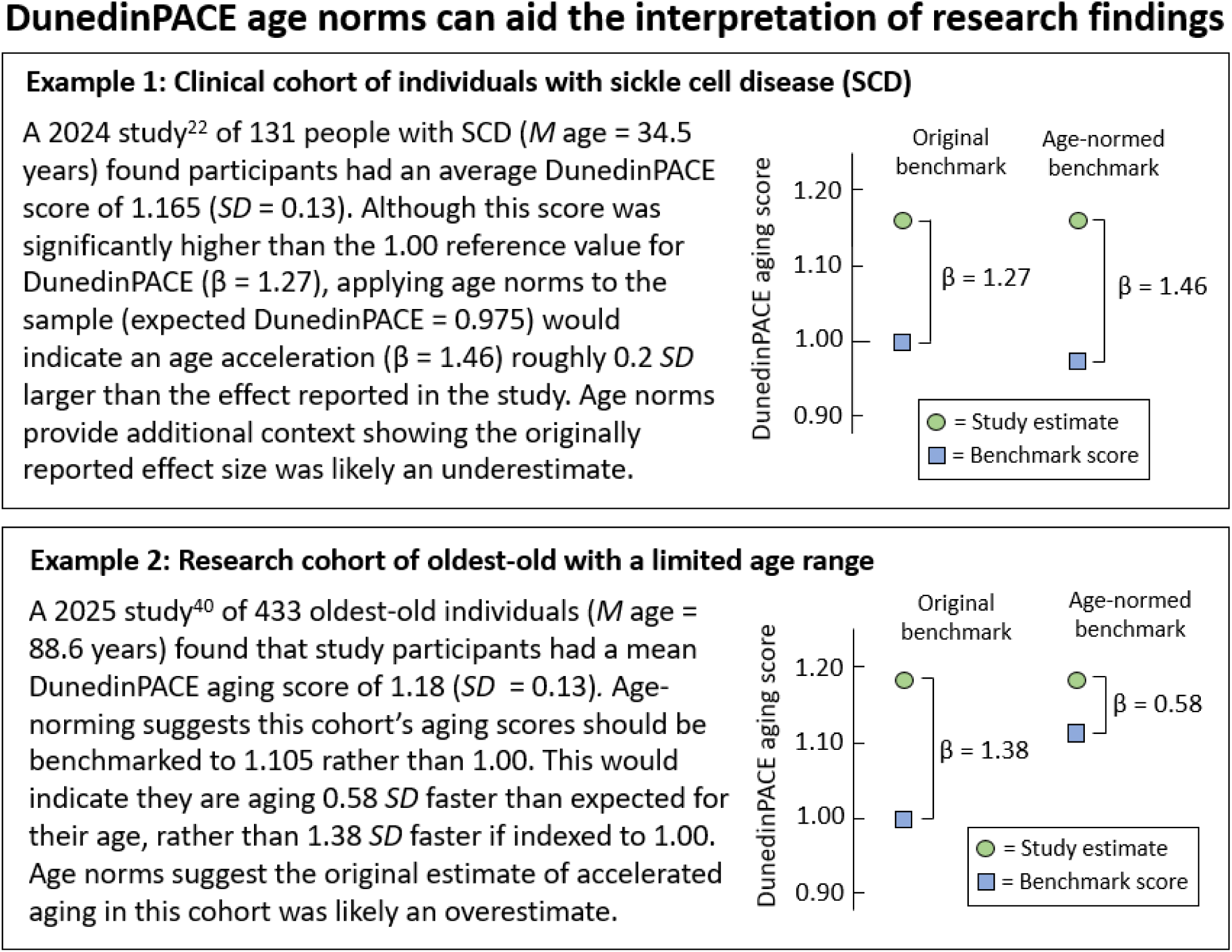
DunedinPACE age norms can aid the interpretation of research findings. Illustration of two examples from recent research studies that show how age norms can assist in the interpretation of DunedinPACE aging score estimates. Estimates of the values used the combined, sex-agnostic age norms **(Supplemental Table 8; Supplemental Figure 1)**, as we did not use individual level data to define the biological sex of the participants. Estimates and 95% confidence intervals were derived using the estimates, *SD*s, and *N*s reported in the two published studies.

## Discussion

We analyzed DNAm data from 37,855 individuals across 11 cohorts to establish age norms for DunedinPACE, the first Pace of Aging epigenetic clock^19^. The findings, age-normed values, online calculator, and updated software reported here increase the interpretability and value of this measure for healthcare and clinical research applications. DunedinPACE was originally benchmarked to the rate of aging observed among 45-year-old New Zealanders. Consistent with evidence that the rate of biological aging accelerates as we grow older, DunedinPACE values tended to be “faster” among older as compared with younger people. This has complicated interpretation of values for young adults (who tend to have values slower than the reference norm of 1.00) and older adults (who tend to have values faster than the reference norm of 1.00). Here, we resolve this complication by generating age-normed reference values across the adult lifespan (ages 20-90). These age norms can be used to generate percentile ranks and confidence intervals for individual scores, similar to how children’s heights and weights are interpreted using population-based references. We created an application (https://tinyurl.com/DunedinPACE-Norms) and code^35^ to support the adoption of these norms in practice and research.

In developing norms, we produced several notable findings. First, Pace of Aging increases linearly across the adult lifespan. Importantly, a linear increase in the rate of aging implies a nonlinear (accelerating) pattern of change in biological age over time. Our results are therefore consistent with observations from the fields of demography, gerontology, and medicine documenting accelerating risk for aging-related disease, functional decline, and mortality across midlife and older age.

Second, the acceleration in Pace of Aging was faster in men compared with women. Although the magnitude of age-dependent acceleration in Pace of Aging is modest overall (2-3% per decade of life), it is roughly twice as fast for men as for women. As with the linear pattern of increase with aging, these findings are consistent with observations across multiple disciplines showing shorter lifespans and earlier onset of aging-related diseases in men compared with women^33^. The relative magnitude of the sex difference highlights the importance of defining sex-specific reference norms for aging biomarkers.

Third, associations between chronological age and DunedinPACE were consistent across cohorts and methods. Our analyses included cohorts spanning multiple countries and continents (North America and Europe), with participants from several genetic ancestries and cultural contexts. The consistency of estimates across cohorts, including differences in the timing and technology of DNAm measurements, suggests that our observations of age patterning are sufficiently robust for clinical translation.

Fourth, our primary results from analysis of between-person age differences were replicated in within-person longitudinal analysis. The within-person estimates were larger than the between-person estimates, though this difference was not statistically significant. These findings confirm that an accelerating rate of aging over the adult lifespan, as measured by DunedinPACE, is a real phenomenon and not an artifact of differences across birth cohorts.

Finally, accounting for the estimated cellular composition of DNA samples attenuated the age patterning of Pace of Aging, but the association remained statistically significant. Aging-related changes in the immune system are well documented, although whether these represent a cofounding influence or a component of the aging phenotype measured by epigenetic clocks remains unresolved^41–42^. Given the profound changes in the immune system that are shaped by environmental exposures in earlier life and the precipitous declines that occur with advancing age, it is challenging to disentangle the influence of age-related immune alterations from aspects of biological aging. Importantly, DunedinPACE predicts clinical outcomes and responses to intervention independent of cell composition^11,19,23–24^. Nevertheless, the observation that variation in immune cell types accounts for a portion of the age patterning of DunedinPACE suggests a need for future study. Analysis of phenotypic Pace of Aging in mixed-age populations with longitudinal data on blood cell composition could help clarify whether our result reflects a technical confound or simply an observation that changes in the immune system are correlated with changes across other systems in the body. Alternatively, these findings could be replicated using Pace of Aging biomarkers that do not rely on blood cell compositions, such as those calculated using structural magnetic resonance imaging^43–44^.

Age norms for epigenetic clocks have implications for future clinical research and practice. Samples used in clinical research can have restricted age ranges, making it challenging to interpret whether differences in Pace of Aging reflect age patterning or the effect of the clinical condition being studied. Using the age norms introduced here, it is now simple to interpret DunedinPACE values without requiring a direct reference population, such as in studies of rare diseases like sickle cell anemia^15^ or exceptional populations, such as centanarians^40^. If future studies measured DunedinPACE in 70-year-old individuals with heart failure or 20-year-old survivors of pediatric cancer, for example, age norms will provide a comparison to the expected rate of aging in same-aged peers.

Age norms support the clinical interpretability of Pace of Aging biomarkers. Until now, formal age norms have not been developed epigenetic clocks, the most commonly used biomarkers of aging. This has limited integration into clinical practice, particularly when compared to the growing body of research linking these measures to important health outcomes^1,2,19,23–24^. The norms developed here, including normative values, percentile ranks, and confidence intervals, provide the needed context to interpret a patient’s Pace of Aging. For example, providers could share how a patient’s score compares to others of their same age and sex, while also accounting for expected age-related increases if DunedinPACE is assessed multiple times in the same patient. The approach we took in this study could also be used to develop age norms for other aging biomarkers to increase their clinical utility.

We acknowledge limitations. Although we used data from 11 independent cohorts spanning multiple nationalities, continents, and race/ethnic groups, they were not inclusive of all human populations. We also did not develop norms for subgroups beyond men and women. Other characteristics might be relevant to interpret rates of aging. For example, genetic ancestry might affect DNAm-based aging biomarkers in ways that lead some populations to look artificially older than others. We have previously established that DunedinPACE scores for non-Hispanic Black veterans do not vary based on their admixture of European and African ancestry^45^. However, care will be needed to monitor application of norms in clinical settings and evaluate if additional population-specific norms are needed. In addition, our age-norms for DunedinPACE cover the adult lifespan, but do not include childhood.

Although there is growing interest in studying biological aging in children, the clinical interpretation of aging values early in the lifespan remains unclear. Finally, our data came from DNAm generated using peripheral blood and 450k and EPIC v1.0 arrays. Illumina has discontinued both of these platforms.

Although our analyses did not detect a difference in age patterning between prior generations of Illumina technology, confirmation that this extends to newer EPIC v2.0 and MSA arrays is needed^40^. Similarly, saliva and dried blood spot samples are increasingly used to generate DNAm data and compute epigenetic clocks. As more data become available, age norms will be needed for clocks calculated from those tissues as well.

## Conclusions

In this study, we used data from 11 independent cohorts, including 43,010 observations across 37,855 individuals, to develop age norms for DunedinPACE, an epigenetic biomarker of the Pace of Aging. These norms included percentile ranks and 95% confidence intervals that can contextualize aging scores for researchers, medical providers, and their patients. Ideally, these norms will support the future scientific and clinical applications of aging biomarkers, particularly as new geroprotective interventions are developed to delay the onset of chronic disease and premature mortality.

## Disclaimer and Role of the Funder/Sponsor

The funders/sponsors had no role in the design and conduct of the study; collection, management, analysis, and interpretation of the data; preparation, review, or approval of the manuscript; and decision to submit the manuscript for publication. The views expressed in this article are those of the authors and do not necessarily reflect the position or policy of the VA, the U.S. government or any other affiliated institution.

## Group information

VA Mid-Atlantic MIRECC Workgroup contributors for this paper include: Patrick S. Calhoun, PhD, Eric Dedert, PhD, Eric B. Elbogen, PhD, Robin A. Hurley, MD, Jason D. Kilts, PhD, Angela Kirby, MS, Scott D. McDonald, PhD, Sarah L. Martindale, Ph.D, Christine E. Marx, MD, MS, Scott D. Moore, MD, PhD, Rajendra A. Morey, MD, MS, Jennifer C. Naylor, PhD, Jared A. Rowland, PhD, Robert D. Shura, PsyD, Cindy Swinkels, PhD, H. Ryan Wagner, PhD.

## Data Sharing

HRS data portal provides access to HRS data (https://hrs.isr.umich.edu/data-products) and NIAGADS (https://dss.niagads.org/datasets/ng00153/). Add Health data can be accessed following study guidance (see https://addhealth.cpc.unc.edu/data). Generation Scotland data access is described here: (https://genscot.ed.ac.uk/for-researchers/access). Data from the Post Deployment Mental Health (PDMH) Study are part of a Veterans Affairs data repository and are available to researchers who request access through the VISN 6 MIRECC and follow the appropriate data access protocols. Data for CARDIA can be accessed through the CARDIA data portal (https://www.cardia.dopm.uab.edu/). WHI data can be accessed through the WHI website (https://www.whi.org/md/working-with-whi-data). All MIDUS data are archived and made publicly available—and are thus shareable—via the University of Michigan Inter-university Consortium of Political and Social Research (ICPSR) or the MIDUS Portal (https://midus.wisc.edu/data-access/). Data for FHS Offspring and Gen3 can be accessed through the FHS website (https://www.framinghamheartstudy.org/fhs-for-researchers/data-available-overview/). Data for SATSA can be accessed through the EMBL-EBL (www.ebi.ac.uk/arrayexpress) and the NACDA (https://www.icpsr.umich.edu/web/NACDA/studies/3843). DNHS data can be accessed through the DNHS website (https://dnhs.unc.edu/)

## References

1. Rutledge J, Oh H, Wyss-Coray T. Measuring biological age using omics data. Nat Rev Genet. 2022;23(12):715–727. doi:10.1038/s41576-022-00511-7

2. Moqri M, Herzog C, Poganik JR, et al. Biomarkers of aging for the identification and evaluation of longevity interventions. Cell. 2023;186(18):3758–3775. doi:10.1016/j.cell.2023.08.003\

3. Biomarkers of Aging Consortium, Herzog CMS, Goeminne LJE, et al. Challenges and recommendations for the translation of biomarkers of aging. Nat Aging. 2024;4(10):1372-1383. doi:10.1038/s43587-024-00683-3

4. Jacques E, Herzog C, Ying K, et al. Invigorating discovery and clinical translation of aging biomarkers. Nat Aging. 2025;5(4):539–543. doi:10.1038/s43587-025-00838-w

5. Ferrucci L, Gonzalez-Freire M, Fabbri E, et al. Measuring biological aging in humans: A quest. Aging Cell. 2020;19(2):e13080. doi:10.1111/acel.13080

6. López-Otín C, Blasco MA, Partridge L, Serrano M, Kroemer G. The hallmarks of aging. Cell. 2013;153(6):1194–1217. doi:10.1016/j.cell.2013.05.039

7. López-Otín C, Blasco MA, Partridge L, Serrano M, Kroemer G. Hallmarks of aging: An expanding universe. Cell. 2023;186(2):243–278. doi:10.1016/j.cell.2022.11.001

8. Moqri M, Herzog C, Poganik JR, et al. Validation of biomarkers of aging. Nat Med. 2024;30(2):360–372. doi:10.1038/s41591-023-02784-9

9. Belsky DW, Baccarelli AA. To promote healthy aging, focus on the environment. Nat Aging. 2023;3(11):1334–1344. doi:10.1038/s43587-023-00518-7

10. Kusters CDJ, Horvath S. Quantification of Epigenetic Aging in Public Health. Annu Rev Public Health. 2025;46(1):91–110. doi:10.1146/annurev-publhealth-060222-015657

11. Waziry R, Ryan CP, Corcoran DL, et al. Effect of long-term caloric restriction on DNA methylation measures of biological aging in healthy adults from the CALERIE trial. Nat Aging. 2023;3(3):248–257. doi:10.1038/s43587-022-00357-y

12. Bischoff-Ferrari HA, Gängler S, Wieczorek M, et al. Individual and additive effects of vitamin D, omega-3 and exercise on DNA methylation clocks of biological aging in older adults from the DO-HEALTH trial. Nat Aging. 2025;5(3):376–385. doi:10.1038/s43587-024-00793-y

13. Li S, Hamaya R, Zhu H, et al. Effects of daily multivitamin-multimineral and cocoa extract supplementation on epigenetic aging clocks in the COSMOS randomized clinical trial. Nat Med, 2026. doi:10.1038/s41591-026-04239-3

14. Horvath S. DNA methylation age of human tissues and cell types. Genome Biol. 2013;14(10):R115. doi:10.1186/gb-2013-14-10-r115

15. Hannum G, Guinney J, Zhao L, et al. Genome-wide methylation profiles reveal quantitative views of human aging rates. Mol Cell. 2013;49(2):359–367. doi:10.1016/j.molcel.2012.10.016

16. Lu AT, Quach A, Wilson JG, et al. DNA methylation GrimAge strongly predicts lifespan and healthspan. Aging. 2019;11(2):303–327. doi:10.18632/aging.101684

17. Levine ME, Lu AT, Quach A, et al. An epigenetic biomarker of aging for lifespan and healthspan. Aging. 2018;10(4):573–591. doi:10.18632/aging.101414

18. Belsky DW, Caspi A, Arseneault L, et al. Quantification of the pace of biological aging in humans through a blood test, the DunedinPoAm DNA methylation algorithm. Elife. 2020;9:e54870. doi:10.7554/eLife.54870

19. Belsky DW, Caspi A, Corcoran DL, et al. DunedinPACE, a DNA methylation biomarker of the pace of aging. Elife. 2022;11:e73420. doi:10.7554/eLife.73420

20. Bourassa KJ, Garrett ME, Caspi A, et al. Posttraumatic stress disorder, trauma, and accelerated biological aging among post-9/11 veterans. Transl Psychiatry. 2024;14(1):4. doi:10.1038/s41398-023-02704-y

21. Crimmins EM, Klopack ET, Kim JK. Generations of epigenetic clocks and their links to socioeconomic status in the Health and Retirement Study. Epigenomics. 2024;16(14):1031–1042. doi:10.1080/17501911.2024.2373682

22. Garrett ME, Le B, Bourassa KJ, et al. Black Americans with Sickle Cell Disease (SCD) demonstrate accelerated epigenetic pace of aging compared to black Americans without SCD. J Gerontol A Biol Sci Med Sci. 2024;79(11):glae230. doi:10.1093/gerona/glae230

23. Bourassa KJ, Anderson L, Woolson S, et al. Accelerated epigenetic aging and prospective morbidity and mortality among U.S. veterans. J Gerontol A Biol Sci Med Sci. 2025;80(7):glaf088. doi:10.1093/gerona/glaf088

24. Bourassa KJ, Dillon KH, Rodriguez RL, et al. Accelerated biological aging and midlife frailty among U.S. military veterans. J Gerontol A Biol Sci Med Sci. 2025;81(1):glaf255. doi:10.1093/gerona/glaf255

25. Finch CE, Crimmins EM. Constant molecular aging rates vs. the exponential acceleration of mortality. Proc Natl Acad Sci. 2016;113(5):1121–1123. doi:10.1073/pnas.1524017113

26. Beard JR, Hanewald K, Si Y, Amuthavalli T J, Moreno-Agostino D. Cohort trends in intrinsic capacity in England and China. Nat Aging. 2025;5(1):87–98. doi:10.1038/s43587-024-00741-w

27. Zenin A, Tsepilov Y, Sharapov S, et al. Identification of 12 genetic loci associated with human healthspan. Commun Biol. 2019;2:41. doi:10.1038/s42003-019-0290-0

28. Shen X, Wang C, Zhou X, et al. Nonlinear dynamics of multi-omics profiles during human aging. Nat Aging. 2024;4(11):1619–1634. doi:10.1038/s43587-024-00692-2

29. Kuo PL, Schrack JA, Levine ME, et al. Longitudinal phenotypic aging metrics in the Baltimore Longitudinal Study of Aging. Nat Aging. 2022;2(7):635–643. doi:10.1038/s43587-022-00243-7

30. Balachandran A, Pei H, Shi Y, et al. Pace of Aging analysis of healthspan and lifespan in older adults in the US and UK. Nat Aging. 2025;5(6):1132–1142. doi:10.1038/s43587-025-00866-6

31. Bell CG, Lowe R, Adams PD, et al. DNA methylation aging clocks: challenges and recommendations. Genome Biol. 2019;20(1):249. Published 2019 Nov 25. doi:10.1186/s13059-019-1824-y

32. Houseman EA, Accomando WP, Koestler DC, Christensen BC, Marsit CJ, Nelson HH, Wiencke JK, Kelsey KT. DNA methylation arrays as surrogate measures of cell mixture distribution. BMC Bioinformat. 2012;13(1):1–6.

33. Salas LA, Koestler DC. FlowSorted.Blood.EPIC: Illumina EPIC data on immunomagnetic sorted peripheral adult blood cells. 2025 doi:10.18129/B9.bioc.FlowSorted.Blood.EPIC, R package version 2.12.0, https://bioconductor.org/packages/FlowSorted.Blood.EPIC

34. Marella WT, Ryan CP, Corcoran D, et al. An epigenetic speedometer to measure Pace of Aging: FraminghamPACE. MedRxiv. 2026. [Preprint]. 10.64898/2026.07.07.26357388

35. Belsky DW. DunedinPACE calculator. 2022. https://github.com/danbelsky/DunedinPACE

36. Austad SN. Why women live longer than men: Sex differences in longevity. Gend Med. 2006;3(2):79–92. doi:10.1016/s1550-8579(06)80198-1

37. Belsky DW, Caspi A, Houts R, et al. Quantification of biological aging in young adults. Proc Natl Acad Sci U S A. 2015;112(30):E4104–E4110. doi:10.1073/pnas.1506264112

38. Moffitt TE, Belsky DW, Danese A, Poulton R, Caspi A. The longitudinal study of aging in human young adults: Knowledge gaps and research agenda. J Gerontol A Biol Sci Med Sci. 2017;72(2):210–215. doi:10.1093/gerona/glw191

39. Mill J, Heijmans BT. From promises to practical strategies in epigenetic epidemiology. Nat Rev Genet. 2013;14(8):585–594. doi:10.1038/nrg3405

40. Tay J, Wang W, Guan L, et al. The Association of Physical Function and Physical Performance With DNA Methylation Clocks in Oldest-Old Living in Singapore-The SG90 Cohort. J Gerontol A Biol Sci Med Sci. 2025;80(4):glaf022. doi:10.1093/gerona/glaf022

41. Jonkman TH, Dekkers KF, Slieker RC, et al. Functional genomics analysis identifies T and NK cell activation as a driver of epigenetic clock progression. Genome Biol. 2022;23(1):24. Published 2022 Jan 14. doi:10.1186/s13059-021-02585-8

42. Jonkman TH; BIOS Consortium, van Zwet EW, Heijmans BT. Probing epigenetic clocks as a rational markers of biological age using blood cell counts. Preprint. medRxiv. 2025;2025.05.12.25327213. Published 2025 May 13. doi:10.1101/2025.05.12.25327213

43. Whitman ET, Elliott ML, Knodt AR, et al. DunedinPACNI estimates the longitudinal Pace of Aging from a single brain image to track health and disease. Nat Aging. 2025;5(8):1619–1636. doi:10.1038/s43587-025-00897-z

44. Whitman ET, Passiatore R, Knodt AR, et al. Replicated evidence for an accelerated rate of whole-body aging in schizophrenia. Psychol Med. 2026;56:e42. Published 2026 Feb 9. doi:10.1017/S003329172610333X

45. Bourassa KJ, Halverson TF, Garrett ME, et al. Demographic characteristics and epigenetic biological aging among post-9/11 veterans: Associations of DunedinPACE with sex, race, and age. Psychiatry Res. 2024;336:115908. doi:10.1016/j.psychres.2024.115908

