## Supplemental materials for "Age norms for DunedinPACE: An epigenetic pace of aging biomarker"

**Supplemental Table 1.** ….…….…….…………..….…………………………………………………………….…………………....……...2

**Supplemental References 1.** ….…….…….…………..….……………………………………………………….…………………...…...5

**Supplemental Table 2.** ….…….…….…………..….…………………………………………………………….…………………....……...6

**Supplemental References 2.** ….…….…….…………..….……………………………………………………….…………………...…...9

**Supplemental Method 1.** ….…….…….…………..….…………….………………………………………….…………………....……...10

**Supplemental References 3.** ….…….…….…………..….……………………………………………………….…………………...…...11

**Supplemental Text 1.** .…….…….…………..….……………………..………………………………………….…………………....……...12

**Supplemental Table 3.** ….…….…….…………..….…………………………………………………………….…………………....……...13

**Supplemental Table 4.** ….…….…….…………..….…………………………………………………………….…………………....……...14

**Supplemental Table 5.** ….…….…….…………..….…………………………………………………………….…………………....……...15

**Supplemental Table 6.** ….…….…….…………..….…………………………………………………………….…………………....……...16

**Supplemental Table 7.** ….…….…….…………..….…………………………………………………………….…………………....……...17

**Supplemental Table 8.** ….…….…….…………..….…………………………………………………………….…………………....……...18

**Supplemental Text 2.** .…….…….…………..….……………………..………………………………………….…………………....……...21

**Supplemental Table 9.** ….…….…….…………..….…………………………………………………………….…………………....……...22

**Supplemental Figure 1.** ….…….…….…………..….…………………………………………………………….…………………....……..23

**Supplemental Table 10.** ..….…….…….…………..….…………………………………………………………….…………………...……24

**Supplemental Method 2.** ….…….…….…………..….…………….………………………………………….…………………....……...25

**Supplemental References 4.** ….…….…….…………..….……………………………………………………….…………………...…...26

**Supplemental Table 1.** *Description of cohorts, characteristics, and measures*

| **Cohort** | **Cohort description** |
| --- | --- |
| Generation Scotland^1^  (*N* = 18,140) | Briefly, the Generation Scotland Scottish Family Health Study is a community- and family-based cohort study of ~24,000 participants across Scotland. During baseline in-clinic assessment between 2006-2011, blood was collected and DNA extracted. The final DNA methylation cohort is majority white (N = 18,142, 97.4%); non-white participants were removed from the data before analysis. For two individuals, recorded age at DNA processing and questionnaire-recorded age was greater than 20%; these two individuals were removed, leaving a final analysis dataset of N = 18,140. Ethical approval for the GS:SFHS study was obtained from the Tayside Committee on Medical Research Ethics (on behalf of the National Health Service), for the GS:3D study was obtained from the Tayside Committee on Medical Research Ethics (on behalf of the National Health Service) and for the GS:21CGH study was obtained from the Scotland A Research Ethics Committee. |
| Add Health^2,3^  (*N* = 4,627) | The National Longitudinal Study of Adolescent to Adult Health (Add Health) is a longitudinal study of a nationally representative cohort of more than 20,000 adolescents in grades 7–12 (aged 12–19) in the United States during the 1994–95 school year. Participants have been followed from adolescence into adulthood across five in-home interviews conducted in 1995, 1996, 2001–2002, 2008–2009, and 2016–2018. Whole blood for DNAm measurement was collected as part of the Wave V biomarker visit and run on the Illumina Infinium Beadchip EPICv1 array^2^. After preprocessing and sample quality control, age, sex, and DNAm for DunedinPACE was available for 4,627 participants. Data can be accessed following study guidance (see https://addhealth.cpc.unc.edu/data) |
| HRS^4^  (*N* = 3,483) | The Health and Retirement Study (HRS) is a nationally representative longitudinal survey of over 37,000 adults aged 50+ across 23,000 U.S. households. Since 1992, data have been collected every two years on health, economic status, cognition, work, retirement, and family connections. The cohort has oversampled African-American and Hispanic households at approximately twice the rate of White households and has maintained strong recruitment and retention of participants. DNA methylation data were generated from blood samples collected during the 2016 Venous Blood Study (VBS) run on the Illumina Infinium Beadchip EPICv1 array. After preprocessing and sample quality control, age, sex, and DNAm for DunedinPACE was available for 3,483 individuals. As a note, a small number (*n* = 77) of individuals were older than 90 and coded as 91, as HRS does not provide exact ages for individuals over 90 years old. Health, demographic, and DNAm data for HRS can be accessed through the HRS data portal (https://hrs.isr.umich.edu/data-products) and NIAGADS (https://dss.niagads.org/datasets/ng00153/). |
| PDMH^5^  (*N* = 2,309) | Veterans were enrolled from 2005 to 2016 in the Veterans Integrated Service Networks 6 (VISN 6) Mental Illness Research, Education, and Clinical Center (MIRECC) Post-Deployment Mental Health Study (PDMH), a multi-site study of veterans who served in the post-9/11 period. A subset of enrolled participants had whole blood drawn and analyzed to generate DNA methylation data, resulting in the final sample of 2,309 veterans. |
| CARDIA^6^  (*N*_inds_=2,023  *N*_obs_=6,214) | The Coronary Artery Risk Development in Young Adults Study (CARDIA) cohort began in 1983 with 5,115 participants and was designed to identify risk factors for future cardiovascular disease. The study enrolled Black and White men and women aged 18 to 30 years from four U.S. cities (Birmingham, Chicago, Minneapolis, Oakland). Whole blood for DNAm was collected across 4 examinations between years 15-30 of the study and run on Illumina Infinium Beadchip EPICv1 array. After preprocessing and sample quality control, age, sex, and DNAm for DunedinPACE was available for 6,214 samples across 2,023 individuals. Health, demographic, and DNAm data for CARDIA can be accessed through the CARDIA data portal (https://www.cardia.dopm.uab.edu/) or dbGaP (Accession: phs001612.v2.p2). |
| WHI^7^  (*N* = 1,845) | The Women’s Health Initiative (WHI) is a long-term national health study focused on postmenopausal women in the United States. The original WHI enrolled 161,808 women aged 50 to 79 between 1993 and 1998 across 40 U.S. clinical centers. WHI includes a randomized Clinical Trial (CT) and an Observational Study (OS), with extensive follow-up through multiple extension studies. Blood samples for DNAm were collected at baseline and run on the Illumina Infinium Beadchip 450k array. After preprocessing and sample quality control, age, sex, and DNAm for DunedinPACE was available for 1,845 individuals. Health, demographic, and DNAm data for WHI can be accessed through the WHI website (https://www.whi.org/md/working-with-whi-data) and dbGaP (Accession: phs001335.v2.p3). |
| MIDUS^8^  (*N* = 1,309) | The Midlife Development in the U.S. (MIDUS) study began in 1995 to investigate the role of behavioral, psychological, and social factors in accounting for age-related variations in health and well-being in a national sample of Americans. Fasting blood draws were obtained from the MIDUS Core sample from 2004 to 2009 and from the MIDUS Refresher sample from 2012 to 2016. |
| FHS Offspring^9^  (*N* = 2,673) | The Framingham Heart Study (FHS) Offspring cohort study began in 1971 with enrollment of 5,124 adult children and spouses of the original 1948 cohort participants from Framingham, Massachusetts. Participants, aged 5–70 years at entry, have undergone repeated medical examinations approximately every four to seven years to study cardiovascular disease (CVD) incidence, risk factors, and family patterns. Our analysis focused on measurements of biological aging from DNA methylation (DNAm) data collected at the eighth follow-up visit and run on the Illumina Infinium Beadchip 450k array. After preprocessing and sample quality control, age, sex, and DNAm for DunedinPACE was available for 2,673 individuals. Health, demographic, and DNAm data for FHS Offspring can be accessed through the FHS website (https://www.framinghamheartstudy.org/fhs-for-researchers/data-available-overview/) and dbGaP (Accession: phs000724.v9.p13) and BioLINCC (Accession: HLB00060025a). |
| DNHS^10^  (*N*_inds_=510  *N*_obs_=846) | The Detroit Neighborhood Health Study (DNHS) is a prospective, longitudinal cohort study of predominantly African American adults living in Detroit, Michigan. Participants were recruited using a dual-frame probability design including landline and cell phone sampling plus mail outreach. Annual structured telephone interviews from 2008 to 2012 assessed neighborhood perceptions, mental and physical health, trauma exposure, and substance use. Biological specimens for genetic and immune biomarker testing were collected from consenting participants. Blood samples for DNAm were collected at multiple timepoints and run on the Illumina Infinium Beadchip EPICv1 array. After preprocessing and sample quality control, age, sex, and DNAm for DunedinPACE was available for 846 samples across 510 individuals. Health, demographic, and DNAm data for DNHS can be accessed through the DNHS website (<https://dnhs.unc.edu/>), the National Addiction and HIV Data Archive Program and dbGaP (Accession: phs000560.v2.p1). |
| FHS Gen3^11^  (*N*=492) | The Framingham Heart Study (FHS) Gen3 sample began in 2002 with enrollment of 4,095 grandchildren of the original 1948 cohort participants from Framingham, Massachusetts. Participants have undergone detailed medical examinations and lifestyle assessments to investigate genetic and environmental factors influencing cardiovascular disease and aging. Blood samples were collected for DNAm at Exam 2 and run on the Illumina Infinium Beadchip EPICv1 array. After preprocessing and sample quality control, age, sex, and DNAm for DunedinPACE was available for 492 individuals. Health, demographic, and DNAm data for FHS Gen3 can be accessed through the FHS website (https://www.framinghamheartstudy.org/fhs-for-researchers/data-available-overview/) and dbGaP (Accession: phs000724.v12.p16) and BioLINCC (Accession: HLB00911223b). |
| SATSA^12^  (*N*_inds_=444  *N*_obs_=1072) | The Swedish Adoption/Twin Study of Aging is a longitudinal cohort study that began in 1984 to examine genetic and environmental influences on aging. The study includes twins separated early in life and raised apart, along with twins raised together as controls. Participants completed repeated questionnaires and a subsample underwent in-person testing on health, cognitive function, and psychosocial factors over multiple waves through 2010. SATSA provides valuable data on how genetics and environment affect aging-related traits, with comprehensive demographic, health, cognitive, and psychosocial measures collected longitudinally. Blood samples were collected for DNAm at multiple timepoints and run on the Illumina Infinium Beadchip 450k array. After preprocessing and sample quality control, age, sex, and DNAm for DunedinPACE was available for 1,072 samples across 444 individuals. Health, demographic, and DNAm data for SATSA can be accessed through the EMBL-EBL ([www.ebi.ac.uk/arrayexpress](https://www.ebi.ac.uk/arrayexpress); accession number E-MTAB-7309 and the NACDA (<https://www.icpsr.umich.edu/web/NACDA/studies/3843>). |

**Supplemental References 1**

1. Walker RM, McCartney DL, Carr K, et al. Data Resource Profile: Whole-Blood DNA Methylation Resource in Generation Scotland (MeGS). *Int J Epidemiol*. 2025;54(4):dyaf091. doi:10.1093/ije/dyaf091
2. Harris, K. M. et al. Cohort Profile: The National Longitudinal Study of Adolescent to Adult Health (Add Health). Int. J. Epidemiol. 48, 1415–1415k (2019).
3. Harris, K. M. *et al.* Sociodemographic and Lifestyle Factors and Epigenetic Aging in US Young Adults: NIMHD Social Epigenomics Program. *JAMA Netw. Open* 7, e2427889 (2024).
4. Sonnega, A. *et al.* Cohort Profile: the Health and Retirement Study. *Int. J. Epidemiol.* 43, 576–585 (2014).
5. Brancu M, Wagner HR, Morey RA, et al. The Post-Deployment Mental Health (PDMH) study and repository: A multi-site study of US Afghanistan and Iraq era veterans. *Int J Methods Psychiatr Res*. 2017;26(3):e1570. doi:10.1002/mpr.1570
6. Lloyd, Jones Donald M. *et al.* The Coronary Artery Risk Development In Young Adults (CARDIA) Study. *JACC* 78, 260–277 (2021).
7. Women’s Health Initiative Study Group. Design of the Women’s Health Initiative clinical trial and observational study. *Control. Clin. Trials* 19, 61–109 (1998).
8. Ryff, Carol D., Seeman, Teresa, and Weinstein, Maxine. Midlife in the United States (MIDUS 2): Biomarker Project, 2004-2009. Inter-university Consortium for Political and Social Research [distributor], 2025-06-18. https://doi.org/10.3886/ICPSR29282.v11
9. Andersson, C., Johnson, A. D., Benjamin, E. J., Levy, D. & Vasan, R. S. 70-year legacy of the Framingham Heart Study. *Nat. Rev. Cardiol.* 16, 687–698 (2019).
10. Aiello, Allison E. (Allison Elizabeth), and Galea, Sandro. Detroit [Michigan] Neighborhood Health Study, 2008-2013. Inter-university Consortium for Political and Social Research [distributor], 2021-10-07. https://doi.org/10.3886/ICPSR37038.v1
11. Splansky, G. L. *et al.* The Third Generation Cohort of the National Heart, Lung, and Blood Institute’s Framingham Heart Study: Design, Recruitment, and Initial Examination. *Am. J. Epidemiol.* 165, 1328–1335 (2007).
12. Pedersen, N. L. *et al.* The Swedish Adoption Twin Study of Aging: An Update. *Acta Genet. Medicae Gemellol. Twin Res.* 40, 7–20 (1991).

**Supplemental Table 2.** *Description of DNA methylation assessment in each cohort*

| **Cohort** | **Chip** | **DNA methylation data generation pipeline** |
| --- | --- | --- |
| Generation Scotland | EPIC v1.0 | DNA methylation profiling was performed on 18,869 individuals in 2016-2021 using the Illumina EPICv1 Beadchip. DNA methylation was assayed in 4 waves. Quality Control and normalization was applied to each wave of data as they were produced. Quality control included of removal of sample outliers on MDS plots, dye-bias and background correction and poor bisulfate conversion. Poorly performing samples and probes were identified via detection p-value > 0.01 in > 0.5% of sites or 1% of samples, and beadcount < 3 in > 5% of samples. Sex was determined from sex-chromosomal DNA methylation patterns and confirmed by self-reported sex. Normalization was performed using dasen from the ‘wateRmelon’ R package^1^. DNA methylation data were provided as M-values; to enable calculation of DunedinPACE values^2^, M-values were converted to Beta-values using the function m2beta in the ‘lumi’ R package^3^ (v2.56.0). |
| Add Health | EPIC v1.0 | Whole blood for DNAm measurement was collected as part of the Wave V biomarker visit and run on the Illumina Infinium Beadchip EPICv1 array. Quality control was carried out at the University of Chapel Hill using using minfi^4^ and ewastools^5^. Samples with signal intensities <10.8 (m) and 9.8 (um), samples showing anomalous beta value density distributions, probes failing bisulfite conversion, samples with mismatched predicted and reported sex, and samples with intermediate genotypes indicating cross-contamination were removed. Probes with polymorphism in the CpG or single basepair extension locus were removed, as were those with detection p-values <0.01 as well as cross-hybridizing probes identified by Pidsley and colleagues^1^. Preprocessing was carried out using the preprocessFunNorm function in minfi^4^. Batch correction was carried out using ComBat^6^. DunedinPACE was derived as per Belsky et al.^2^ |
| HRS | EPIC v1.0 | DNA methylation assays were done on a subsample of 4,104 participants from the 2016 Venous Blood Study. DNA methylation data are based on assays done using the Infinium Methylation EPIC BeadChip v1.0 at the University of Minnesota. Preprocessing and normalization of DNA methylation data were conducted at the Robert N. Butler Columbia Aging Center Geroscience Computational Core (GCC) in R (v 4.4.1) using minfi^4^, wateRmelon^1^, and ewastools^5^. IDATs were matched to phenotype information and partitioned into 5 chunks of roughly equal size. Samples with the following criteria were removed: samples methylated or unmethylated signal intensity <10.5; samples with >5% of probes with fewer than 5 beads; samples with bisulfite conversion efficiency <80%; samples with an average detection p-value ≥0.05; samples with irregular clustering of SNP-based probes. Additionally, 17 metrics implemented in the ewastools package^10^ were applied. Normalization was performed using the preprocessNoob function in the minfi^4^ package. DunedinPACE was derived as per Belsky et al.^2^ Cell composition was estimated using the minfi estimateCellCounts function^7^. |
| PDMH | 450k, EPIC v1.0 | Whole blood was collected at the baseline PDMH assessment and analyzed using the Infinium HumanMethylation450 or MethylationEPIC v1.0 Beadchip (Illumina Inc., San Diego, CA) to derive DNAm data. Internal replicates were checked for consistency using single nucleotide polymorphisms on each array. Quality control was performed using the minfi^4^ and ChAMP R packages^8^. Probe quality control and data normalization were performed within each batch using the R package wateRmelon^1^. Raw beta values were normalized using the dasen approach^1^, and batch and chip adjustments were completed using ComBat in the R package sva^6^. White blood cell composition was estimated using the Houseman approach^9^. |
| CARDIA | EPIC v1.0 | Whole blood for DNAm was collected across 4 examinations between years 15-30 of the study and run on Illumina Infinium Beadchip EPICv1 array. Preprocessing and normalization of DNA methylation data were conducted at the Robert N. Butler Columbia Aging Center Geroscience Computational Core (GCC) in R (v 4.4.1) using minfi^4^, wateRmelon^1^, and ewastools^5^. IDATs were matched to phenotype information and partitioned into 8 chunks of roughly equal size. When present, replicate and longitudinal samples from the same individual were kept together in the same chunk. Samples with the following criteria were removed: samples methylated or unmethylated signal intensity <10.5; samples with >5% of probes with fewer than 5 beads; samples with bisulfite conversion efficiency <80%; samples with an average detection p-value ≥0.05; samples with irregular clustering of SNP-based probes. Additionally, 17 metrics implemented in the ewastools package^10^ were applied. Normalization was performed using the preprocessNoob function in the minfi^4^ package. DunedinPACE was derived as per Belsky et al.^2^. Cell composition was estimated using the minfi estimateCellCounts2 function and the Salas extended reference panel^7^. |
| WHI | 450k | Blood samples for DNAm were collected at baseline and run on the Illumina Infinium Beadchip 450k array. Preprocessing and normalization of DNA methylation data were conducted at the Robert N. Butler Columbia Aging Center Geroscience Computational Core (GCC) in R using minfi^4^, wateRmelon^1^, and ewastools^5^. IDATs were matched to phenotype information and samples with the following criteria were removed: samples methylated or unmethylated signal intensity <10.5; samples with >5% of probes with fewer than 5 beads; samples with bisulfite conversion efficiency <80%; samples with an average detection p-value ≥0.05; samples with irregular clustering of SNP-based probes. Additionally, 17 metrics implemented in the ewastools package^5^ were applied. Normalization was performed using the preprocessNoob function in the minfi^4^ package. DunedinPACE was derived as per Belsky et al^2^. Cell composition was estimated using the minfi estimateCellCounts2 function and the Salas extended reference panel^7^. |
| MIDUS | EPIC v1.0 | DNA was extracted from whole blood collected using a BD Vacutainer Tube with EDTA anticoagulant and frozen in storage. In 2019, DNA methylation profiling was conducted on the whole blood DNA samples from both the Core and Refresher samples. Whole blood was tested for suitable DNA yield and DNA integrity, and had methylation assessed using Illumina Methylation EPIC microarrays. The methylation values were normalized to control for technical sources of variance using the noob function in the R minfi package^4^, registered onto the list of CpG sites assayed on the Illumina Methylation 450K microarray, screened using standard quality control metrics for DNAm array data, and scored using previously published algorithms^2^. Notably, MIDUS does not release CpG level data, all aging score values are already calculated in the final dataset, which precludes white blood cell estimates derived from DNAm, as they are not provided by MIDUS. |
| FHS Offspring | EPIC v1.0 | Whole blood for DNA methylation (DNAm) was collected at the eighth follow-up visit and run on the Illumina Infinium Beadchip 450k array. Preprocessing and normalization of DNA methylation data were conducted at the Robert N. Butler Columbia Aging Center Geroscience Computational Core (GCC) in R using minfi^4^, wateRmelon^1^, and ewastools^5^. IDATs were matched to phenotype information and samples with the following criteria were removed: samples methylated or unmethylated signal intensity <10.5; samples with >5% of probes with fewer than 5 beads; samples with bisulfite conversion efficiency <80%; samples with an average detection p-value ≥0.05; samples with irregular clustering of SNP-based probes. Additionally, 17 metrics implemented in the ewastools package^5^ were applied. Normalization was performed using the preprocessNoob function in the minfi^4^ package. DunedinPACE was derived as per Belsky et al^2^. Cell composition was estimated using the minfi estimateCellCounts2 function and the Salas extended reference panel^7^. |
| DNHS | EPIC v1.0 | Blood samples for DNAm were collected at multiple timepoints and run on the Illumina Infinium Beadchip EPICv1 array. Preprocessing was conducted using ewastools^5^. Samples with probe detection call rates lower than 90% and average intensity values that were either less than 50% of the overall sample mean or below 2000 arbitrary units (AU) were excluded. Probes with detection *p*-values > 0.01 were considered low quality and treated as missing. Probes that were missing in > 10% of the samples within the studies and were cross-hybridizing were removed. Data was normalized using single-sample Noob (ssNoob) implemented in R package minfi^4^. *ComBat* was used to account for batch effects of chip and position^6^. DunedinPACE was derived as per Belsky et al^2^. Blood-cell was estimated using the robust partial correlation (RPC) method in Epidish with a reference data specific to EPIC array. Code for sample and probe quality control can be found at https://github.com/PGC-PTSD-EWAS/EPIC_QC/tree/main. |
| FHS 3^rd^ Gen | EPIC v1.0 | Blood samples were collected for DNAm at Exam 2 and run on the Illumina Infinium Beadchip EPICv1 array. Preprocessing and normalization of DNA methylation data were conducted at the Robert N. Butler Columbia Aging Center Geroscience Computational Core (GCC) in R using minfi^4^, wateRmelon^1^, and ewastools^5^. IDATs were matched to phenotype information and samples with the following criteria were removed: samples methylated or unmethylated signal intensity <10.5; samples with >5% of probes with fewer than 5 beads; samples with bisulfite conversion efficiency <80%; samples with an average detection p-value ≥0.05; samples with irregular clustering of SNP-based probes. Additionally, 17 metrics implemented in the ewastools package^5^ were applied. Normalization was performed using the preprocessNoob function in the minfi^4^ package. DunedinPACE was derived as per Belsky et al^2^. Cell composition was estimated using the minfi estimateCellCounts2 function and the Salas extended reference panel^7^. |
| SATSA | 450k | Blood samples were collected for DNAm at multiple timepoints and run on the Illumina Infinium Beadchip 450k array. Preprocessing and normalization of DNA methylation data were conducted at the Robert N. Butler Columbia Aging Center Geroscience Computational Core (GCC) in R using minfi^4^, wateRmelon^1^, and ewastools^5^. Sample quality controls measures included examining the signal intensities (<10.5), number of probes with <5 beads (>5%), samples with low bisulfite conversion efficiency (<80%), samples with average detection p-value ≥0.05; samples with irregular clustering of SNP-based probes, and 17 metrics implemented in the ewastools package^5^. A large number of samples (596) failed sample quality control metrics. To maximize data availability, this analysis retained all samples, including those that failed quality control metrics. Normalization was performed using the preprocessNoob function in the minfi^4^ package. DunedinPACE was derived as per Belsky et al^2^. Cell composition was estimated using the minfi estimateCellCounts2 function and the Salas extended reference panel^7^. |

**Supplemental References 2**

1. Pidsley R, Y Wong CC, Volta M, Lunnon K, Mill J, Schalkwyk LC. A data-driven approach to preprocessing Illumina 450K methylation array data. *BMC Genomics*. 2013;14:293. Published 2013 May 1. doi:10.1186/1471-2164-14-293
2. Belsky DW, Caspi A, Corcoran DL, et al. DunedinPACE, a DNA methylation biomarker of the pace of aging. *Elife*. 2022;11:e73420. Published 2022 Jan 14. doi:10.7554/eLife.73420
3. Du P, Kibbe WA, Lin SM. lumi: a pipeline for processing Illumina microarray. *Bioinformatics*. 2008;24(13):1547-1548. doi:10.1093/bioinformatics/btn224
4. Aryee MJ, Jaffe AE, Corrada-Bravo H, et al. Minfi: a flexible and comprehensive Bioconductor package for the analysis of Infinium DNA methylation microarrays. *Bioinformatics*. 2014;30(10):1363-1369. doi:10.1093/bioinformatics/btu049
5. Heiss JA, Just AC. Identifying mislabeled and contaminated DNA methylation microarray data: an extended quality control toolset with examples from GEO. *Clin Epigenetics*. 2018;10:73. Published 2018 Jun 1. doi:10.1186/s13148-018-0504-1
6. Leek JT, Johnson WE, Parker HS, Jaffe AE, Storey JD. The sva package for removing batch effects and other unwanted variation in high-throughput experiments. *Bioinformatics*. 2012;28(6):882-883. doi:10.1093/bioinformatics/bts034
7. Salas LA, Zhang Z, Koestler DC, et al. Enhanced cell deconvolution of peripheral blood using DNA methylation for high-resolution immune profiling. *Nat Commun*. 2022;13(1):761. Published 2022 Feb 9. doi:10.1038/s41467-021-27864-7
8. Morris TJ, Butcher LM, Feber A, Teschendorff AE, Chakravarthy AR, Wojdacz TK, Beck S. ChAMP: 450k Chip Analysis Methylation Pipeline. *Bioinformatics*. 2014;30(3):428-430. doi:10.1093/bioinformatics/btt684
9. Houseman EA, Accomando WP, Koestler DC, Christensen BC, Marsit CJ, Nelson HH, et al. DNA methylation arrays as surrogate measures of cell mixture distribution. *BMC Bioinformat*. 2012;13(1):1-6.

**Supplemental Method 1.**

Most of the studies we examined included a single observation per individual. For these studies, we estimated the association between chronological age and DunedinPACE using linear regression models and ordinary least squares (OLS). Linear regression results were reported for Generation Scotland, but we also conducted panel linear models (PLM) and generalized estimating equations (GEE) to account for clustering within family, which was present in the data. Accounting for clustering by family using PLM and GEE did not substantively alter any of the results, so we chose to report the linear regression results throughout the main text. A subset of studies (SATSA, CARDIA, DNHS) included more than one observation per individual. For these studies, we modeled associations using GEE with an unstructured working correlation to account for repeated measures within individuals. For OLS models, we report the conventional coefficient of determination (R²). For GEE models with multiple observations, we removed all but the first observation for each individual and report the conventional coefficient of determination (R²) from the OLS model summary fit on these observations.

For OLS models, we computed the Bayesian Information Criterion (BIC) using the standard maximum-likelihood formulation implemented in stats::BIC function. For GEE models, we used a custom code pipeline that approximates BIC using a quasi-likelihood for Gaussian outcomes^1^. For studies with repeated measures per participant, we also estimated within-person associations between changes in DunedinPACE and changes in chronological age using fixed-effects regression models (via fixest::feols in R). Within each cohort, age at measurement was centered within person to ensure the model estimated within-person rather than between-person effects. We fit linear, quadratic, and cubic models of age, including individual fixed effects^2^ to account for all time-invariant covariates (e.g., sex, baseline characteristics, and genetic factors). Standard errors were clustered at the individual level, and model fit was quantified manually using the within-person R. In contrast to GEE, which estimate population-averaged (marginal) effects while accounting for within-subject correlation, fixed-effects models estimate subject-specific effects that explicitly remove between-person variation. This makes them well suited for assessing within-individual trajectories over time

**Supplemental References 3**

1. Pan, W. (2001). Akaike’s information criterion in generalized estimating equations. Biometrics, 57(1), 120–125. https://doi.org/10.1111/j.0006-341X.2001.00120.x
2. Gunasekara, F. I., Richardson, K., Carter, K., & Blakely, T. (2014). Fixed effects analysis of repeated measures data. International Journal of Epidemiology, 43(1), 264–269. https://doi.org/10.1093/ije/dyt221

**Supplemental Text 1.**

We examined whether the association between chronological age and DunedinPACE was non-linear using three approaches applied in the full cohorts, as well in models stratified by sex.

First, we meta-analyzed whether quadratic or cubic coefficients were associated with DunedinPACE when added to a linear coefficient. Neither quadratic (*B* = -0.0213, 95% CI [-0.0527, 0.0102], *p* = .185) nor cubic age coefficients (*B* = 0.0126, 95% CI [-0.0128, 0.0379], *p* = .332) were associated with DunedinPACE. These results were similar for both men and women (all *p*s > .30). Full results for each cohort are reported in **Supplemental Table 3**.

Second, we examined whether quadratic and cubic age coefficients improved model fit, assessed using BIC, with decreases (ΔBIC < -2) representing improvement for the nested models. Only 2 of the 11 cohorts showed improvement in model fit when adding a quadratic coefficient (mean ΔBIC across cohorts = +1.24), with 2 separate cohorts showing improvement in model fit when adding a cubic coefficients (mean ΔBIC across cohorts = +3.04). Said differently, 90.9% of the nested models (40 of 44) showed no improvement when adding a non-linear coefficient. Results were similar for men and women when modeled independently. Full results for each cohort are reported in **Supplemental Table 4**).

Third, we tested whether non-linear age coefficients explained additional variance in DunedinPACE. Quadratic (mean Δ*R^2^* across cohorts = 0.2%) and cubic functions (mean Δ*R^2^* = 0.1%) explained a small proportion of additional variance compared to a linear model (mean variance explained across cohorts = 5.0%). Full results for each cohort are reported in **Supplemental Table 5**. Said differently, a linear model accounted for 96.2% of the variance explained by a model including a quadratic coefficient, and 94.3% of the variance explained by a model including a cubic coefficient. This pattern of results was similar among men (quadratic Δ*R^2^* = 0.3%; cubic Δ*R^2^* = 0.2%) and women (quadratic Δ*R^2^* = 0.1%; cubic Δ*R^2^* = 0.3%)**.**

**Supplemental Table 3.** *Meta-analyzing the association of quadratic and cubic age coefficients with DunedinPACE*

| **Full cohort** | | | | | | |
| --- | --- | --- | --- | --- | --- | --- |
|  |  | **Quadratic** | | | **Cubic** | |
| **Cohort** | **Obs.** | ***B*** | **95% CI** | ***B*** | | **95% CI** |
| HRS | 3,483 | 0.0164 | -0.0316, 0.0644 | -0.0233 | | -0.0679, 0.0213 |
| CARDIA | 6,214 | -0.0246 | -0.0654, 0.0162 | 0.0908 | | 0.0433, 0.1383 |
| WHI | 1,845 | -0.0578 | -0.1620, 0.0465 | 0.0537 | | -0.0804, 0.1878 |
| FHS Offspring | 2,673 | -0.0051 | -0.0521, 0.0419 | -0.0648 | | -0.1012, -0.0283 |
| FHS 3rdGen | 492 | -0.0517 | -0.1275, 0.0241 | 0.0427 | | -0.0079, 0.0933 |
| DNHS | 846 | -0.0720 | -0.0999, -0.0441 | 0.0144 | | 0.0003, 0.0286 |
| SATSA | 1,072 | 0.1383 | 0.0532, 0.2234 | 0.0280 | | -0.0432, 0.0992 |
| Add Health | 4,627 | 0.2067 | -0.6624, 1.0757 | 3.2928* | | -0.0321, 6.6179 |
| MIDUS | 1,309 | -0.0322 | -0.0718, 0.0074 | 0.0154 | | -0.0093, 0.0401 |
| PDMH | 2,309 | -0.0793 | -0.1213, -0.0374 | 0.0073 | | -0.0273, 0.0419 |
| Generation Scotland | 18,140 | -0.0084 | -0.0153, -0.0015 | 0.0087 | | 0.0052, 0.0122 |
| **Meta-analyzed results** | | **-0.0213** | **-0.0527, 0.0102** | **0.0126** | | **-0.0128, 0.0379** |
| **Women** | | | | | | |
| **Cohort** | **Obs.** | ***B*** | **95% CI** | ***B*** | | **95% CI** |
| HRS | 2,048 | 0.0324 | -0.0309, 0.0958 | -0.0124 | | -0.0707, 0.0458 |
| CARDIA | 3,669 | -0.0866 | -0.1393, -0.0338 | 0.1046 | | 0.0433, 0.1659 |
| WHI | 1,845 | -0.0578 | -0.1620, 0.0465 | 0.0537 | | -0.0804, 0.1878 |
| FHS Offspring | 1,457 | 0.0645 | 0.0019, 0.1271 | -0.0785 | | -0.1252, -0.0318 |
| FHS 3rdGen | 262 | -0.0829 | -0.1960, 0.0303 | 0.0877 | | 0.0061, 0.1693 |
| DNHS | 509 | -0.0602 | -0.0997, -0.0207 | 0.0029 | | -0.0143, 0.0201 |
| SATSA | 647 | 0.1094 | 0.0123, 0.2065 | -0.0049 | | -0.0896, 0.0798 |
| Add Health | 2,786 | 0.8210 | -0.3688, 2.0108 | 3.7371* | | -1.0881, 8.0946 |
| MIDUS | 730 | -0.0418 | -0.0962, 0.0126 | 0.0164 | | -0.0169, 0.0497 |
| PDMH | 491 | -0.1080 | -0.2004, -0.0156 | -0.1410 | | -0.2408, -0.0412 |
| Generation Scotland | 10,673 | -0.0030 | -0.0120, 0.0060 | 0.0170 | | 0.0125, 0.0215 |
| **Meta-analyzed results** | | **-0.0213** | **-0.0618, 0.0191** | **0.0051** | | **-0.0348, 0.0450** |
| **Men** | | | | | | |
| **Cohort** | **Obs.** | ***B*** | **95% CI** | ***B*** | | **95% CI** |
| HRS | 1,435 | 0.0001 | -0.0729, 0.0730 | -0.0488 | | -0.1194, 0.0217 |
| CARDIA | 2,545 | 0.0516 | -0.0118, 0.1150 | 0.0613 | | -0.0148, 0.1373 |
| WHI | 0 | — | — | — | | — |
| FHS Offspring | 1,216 | -0.0857 | -0.1539, -0.0175 | -0.0372 | | -0.0929, 0.0185 |
| FHS 3rdGen | 230 | -0.0352 | -0.1348, 0.0645 | 0.0026 | | -0.0625, 0.0678 |
| DNHS | 337 | -0.0800 | -0.1205, -0.0395 | 0.0246 | | 0.0014, 0.0478 |
| SATSA | 425 | 0.2331 | 0.0778, 0.3883 | 0.1297 | | 0.0056, 0.2538 |
| Add Health | 1,841 | -0.3377 | -1.5951, 0.9197 | 3.4919* | | -1.1108, 8.0946 |
| MIDUS | 579 | -0.0310 | -0.0898, 0.0278 | 0.0135 | | -0.0226, 0.0496 |
| PDMH | 1,818 | -0.0667 | -0.1131, -0.0203 | 0.0025 | | -0.0289, 0.0339 |
| Generation Scotland | 7,467 | -0.0083 | -0.0191, 0.0025 | -0.0026 | | -0.0083, 0.0031 |
| **Meta-analyzed results** | | **-0.0201** | **-0.0608, 0.0205** | **0.0054** | | **-0.0085, 0.0193** |
| *Note*: The linear model includes only the association of DunedinPACE and (linear) age. Quadratic adds a quadratic term to the model, and cubic add a cubic term to the model. Quadratic and cubic age were divided by 1,000 and 10,000 respectively to improve the legibility of the estimates. Obs. = observations. * Estimates for Add Health were large in magnitude due to a restriction in range for age, however, all substantive results of the meta-analyses are unchanged if excluding the Add Health cohort in leave-one-out analyses. | | | | | | |

**Supplemental Table 4.** *Testing the change in model fit when adding non-linear age terms*

| **Full cohort** | | | | | | | |
| --- | --- | --- | --- | --- | --- | --- | --- |
|  | **Linear** | | **Quadratic** | | | **Cubic** | |
| **Cohort** | **Obs.** | **BIC** | **BIC** | **ΔBIC** | **BIC** | | **ΔBIC** |
| HRS | 3,483 | -4003.61 | -3995.91 | +7.71 | -3988.81 | | +7.10 |
| CARDIA | 6,214 | -7410.91 | -7401.74 | +9.17 | -7393.81 | | +7.93 |
| WHI | 1,845 | -2481.20 | -2474.87 | +6.34 | -2467.96 | | +6.90 |
| FHS Offspring | 2,673 | -3617.79 | -3609.94 | +7.85 | **-3614.18** | | **-4.24** |
| FHS 3rdGen | 492 | -774.26 | -769.87 | +4.39 | -766.43 | | +3.44 |
| DNHS | 846 | -1072.19 | **-1104.26** | **-32.07** | -1103.85 | | +0.41 |
| SATSA | 1,072 | -812.73 | -813.51 | -0.78 | -806.79 | | +6.72 |
| Add Health | 4,627 | -5501.08 | -5492.85 | +8.22 | -5488.18 | | +4.67 |
| MIDUS | 1,309 | -1503.06 | -1498.43 | +4.63 | -1492.76 | | +5.67 |
| PDMH | 2,309 | -3612.16 | **-3618.10** | **-5.94** | -3610.53 | | +7.57 |
| Generation Scotland | 18,140 | -24745.59 | -24741.41 | +4.18 | **-24754.09** | | **-12.68** |
| ***N* models improved (mean Δ)** | |  | **2 of 11 (+1.24)** | | **2 of 11 (+3.04)** | | |
| **Women** | | | | | | | |
| **Cohort** | **Obs.** | **BIC** | **BIC** | **ΔBIC** | **BIC** | | **ΔBIC** |
| HRS | 2,048 | -2179.00 | -2172.39 | +6.62 | -2164.94 | | +7.45 |
| CARDIA | 3,669 | -4179.08 | -4170.53 | +8.56 | -4163.51 | | +7.02 |
| WHI | 1,845 | -2481.20 | -2474.87 | +6.34 | -2467.96 | | +6.90 |
| FHS Offspring | 1,457 | -2023.47 | -2020.28 | +3.19 | **-2023.87** | | **-3.59** |
| FHS 3rdGen | 262 | -409.03 | -405.56 | +3.47 | -404.50 | | +1.06 |
| DNHS | 509 | -632.59 | **-640.97** | **-8.38** | -634.82 | | +6.14 |
| SATSA | 647 | -486.45 | -483.44 | +3.01 | -477.02 | | +6.43 |
| Add Health | 2,786 | -3267.26 | -3261.16 | +6.10 | -3255.54 | | +5.62 |
| MIDUS | 730 | -820.23 | -815.9 | +4.33 | -810.25 | | +5.65 |
| PDMH | 491 | -756.75 | -755.78 | +0.97 | -749.71 | | +6.07 |
| Generation Scotland | 10,673 | -14,392.35 | -14,383.53 | +8.82 | **-14,426.72** | | **-43.19** |
| ***N* models improved (mean Δ)** | |  | **1 of 11 (+3.91)** | | **2 of 11 (+0.51)** | | |
| **Men** | | | | | | | |
| **Cohort** | **Obs.** | **BIC** | **BIC** | **ΔBIC** | **BIC** | | **ΔBIC** |
| HRS | 1,435 | -1861.45 | -1854.18 | +7.27 | -1848.76 | | +5.42 |
| CARDIA | 2,545 | -3250.96 | -3243.21 | +7.75 | -3235.21 | | +7.99 |
| WHI | 0 | — | — | — | — | | — |
| FHS Offspring | 1,216 | -1676.00 | -1674.97 | +1.03 | -1669.58 | | +5.38 |
| FHS 3rdGen | 230 | -367.39 | -362.44 | +4.95 | -357.01 | | +5.43 |
| DNHS | 337 | -434.24 | **-450.33** | **-16.09** | **-453.72** | | **-3.39** |
| SATSA | 425 | -329.67 | -331.35 | -1.68 | -328.53 | | +2.82 |
| Add Health | 1,841 | -2282.21 | -2274.97 | +7.24 | -2269.67 | | +5.30 |
| MIDUS | 579 | -689.12 | -683.90 | +5.22 | -678.08 | | +5.82 |
| PDMH | 1,818 | -2897.39 | -2897.80 | -0.41 | -2890.31 | | +7.49 |
| Generation Scotland | 7,467 | -10630.50 | -10623.85 | +6.65 | -10615.76 | | +8.09 |
| ***N* models improved (mean Δ)** | |  | **1 of 11 (+2.19)** | | **1 of 11 (+5.04)** | | |
| *Note*: The linear model includes the association of DunedinPACE and age (linear) only. Quadratic adds a quadratic term to the model, and cubic add a third cubic term to the model. Bolded BIC values represent an improvement in model fit (Δ < -2). BIC = Bayesian Information Criteria, Obs. = observations. | | | | | | | |

**Supplemental Table 5.** *Testing the added variance in DunedinPACE explained when adding non-linear age terms*

| **Cohort** | **Linear** | | **Quadratic** | | | **Cubic** | |
| --- | --- | --- | --- | --- | --- | --- | --- |
| **All participants** | **Obs.** | ***R^2^*** | ***R^2^*** | **Δ*R^2^*** | ***R^2^*** | | **Δ*R^2^*** |
| HRS | 3,483 | 1.1% | 1.1% | 0.0% | 1.1% | | 0.1% |
| CARDIA | 6,214 | 1.9% | 2.3% | 0.4% | 2.3% | | 0.0% |
| WHI | 1,845 | 0.5% | 0.6% | 0.1% | 0.6% | | 0.0% |
| FHS Offspring | 2,673 | 8.7% | 8.7% | 0.0% | 9.1% | | 0.4% |
| FHS 3rdGen | 492 | 3.2% | 3.5% | 0.3% | 4.1% | | 0.6% |
| DNHS | 846 | 4.8% | 8.9% | 4.1% | 9.5% | | 0.6% |
| SATSA | 1,072 | 5.4% | 6.1% | 0.7% | 6.1% | | 0.0% |
| Add Health | 4,627 | 0.7% | 0.7% | 0.0% | 0.8% | | 0.1% |
| MIDUS | 1,309 | 3.3% | 3.5% | 0.2% | 3.6% | | 0.1% |
| PDMH | 2,309 | 5.5% | 6.1% | 0.6% | 6.1% | | 0.0% |
| Generation Scotland | 18,140 | 7.9% | 7.9% | 0.0% | 8.0% | | 0.1% |
| **Weighted average** |  | 5.0% | 5.2% | 0.2% | 5.3% | | 0.1% |
| **Women** | **Obs.** | ***R^2^*** | ***R^2^*** | **Δ*R^2^*** | ***R^2^*** | | **Δ*R^2^*** |
| HRS | 2,048 | 0.7% | 0.7% | 0.0% | 0.8% | | 0.1% |
| CARDIA | 3,669 | 2.1% | 2.2% | 0.1% | 2.2% | | 0.0% |
| WHI | 1,845 | 0.5% | 0.6% | 0.1% | 0.6% | | 0.1% |
| FHS Offspring | 1,457 | 8.5% | 8.7% | 0.2% | 9.4% | | 0.7% |
| FHS 3rdGen | 262 | 0.3% | 1.1% | 0.8% | 2.8% | | 1.7% |
| DNHS | 509 | 1.7% | 4.2% | 2.5% | 4.4% | | 0.2% |
| SATSA | 647 | 6.0% | 6.4% | 0.4% | 6.9% | | 0.5% |
| Add Health | 2,786 | 0.7% | 0.7% | 0.0% | 0.8% | | 0.1% |
| MIDUS | 730 | 0.3% | 0.6% | 0.3% | 0.7% | | 0.1% |
| PDMH | 491 | 2.9% | 4.0% | 1.1% | 4.0% | | 0.0% |
| Generation Scotland | 10,673 | 3.4% | 3.4% | 0.0% | 3.8% | | 0.4% |
| **Weighted average** | | 2.7% | 2.8% | 0.1% | 3.1% | | 0.3% |
| **Men** | **Obs.** | ***R^2^*** | ***R^2^*** | **Δ*R^2^*** | ***R^2^*** | | **Δ*R^2^*** |
| HRS | 1,435 | 1.6% | 1.6% | 0.0% | 1.7% | | 0.1% |
| CARDIA | 2,545 | 3.9% | 4.6% | 0.7% | 4.7% | | 0.1% |
| WHI | 0 | — | — | — | — | | — |
| FHS Offspring | 1,216 | 10.0% | 10.4% | 0.4% | 11.0% | | 0.6% |
| FHS 3rdGen | 230 | 8.8% | 9.0% | 0.2% | 9.0% | | 0.0% |
| DNHS | 337 | 10.6% | 15.8% | 5.2% | 16.6% | | 0.8% |
| SATSA | 425 | 6.9% | 8.2% | 1.3% | 9.3% | | 1.1% |
| Add Health | 1,841 | 1.2% | 1.2% | 0.0% | 1.3% | | 0.1% |
| MIDUS | 579 | 10.5% | 10.7% | 0.2% | 10.8% | | 0.1% |
| PDMH | 1,818 | 6.5% | 6.9% | 0.4% | 6.9% | | 0.0% |
| Generation Scotland | 7,467 | 17.3% | 17.4% | 0.1% | 17.4% | | 0.0% |
| **Weighted average** | | 10.2% | 10.5% | 0.3% | 10.7% | | 0.2% |
| *Note*: The linear model includes the association of DunedinPACE and age (linear) only. Quadratic adds a quadratic term to the model, and cubic add a third cubic term to the model. R^2^ represented the total variance explained by the model, Δ*R^2^* represents the increase in variance explained by adding the quadratic and then cubic term. Weighted average accounts for sample size. Obs. = observations. | | | | | | | |

**Supplemental Table 6.** *Testing the association of chronological age and DunedinPACE using longitudinal data*

|  |  | **Cross-sectional association** | | | **Longitudinal association** | |
| --- | --- | --- | --- | --- | --- | --- |
| **Cohort** | ***N*** | **Obs.** | ***B*** | **95% CI** | ***B*** | **95% CI** |
| CARDIA | 2,023 | 6,214 | 0.0017 | 0.0013, 0.0021 | 0.0021 | 0.0017, 0.0024 |
| DNHS | 510 | 846 | 0.0019 | 0.0012, 0.0025 | 0.0065 | 0.0036, 0.0093 |
| SATSA | 444 | 1,072 | 0.0042 | 0.0030, 0.0054 | 0.0042 | 0.0025, 0.0059 |
| **Meta-analyzed results** | | | 0.0025 | 0.0010, 0.0040 | 0.0039 | 0.0015, 0.0064 |
| *Note*: There were an average of 3.07 (*SD* = 0.88) DNAm assessments in CARDIA, 2.41 (*SD* = 1.31) assessments in SATSA, and 1.66 (*SD* = 0.86) assessments in DNHS. The time between DNAm assessments averaged 5.03 years (*SD* = 0.95) in CARDIA, 5.40 years (*SD* = 1.99) in SATSA, and 2.19 years (*SD* = 1.01) in DNHS. Meta-analyzed results were generated using a random effects model. *N* represents the number of unique individuals, Obs. represents the number of observations across those individuals. | | | | | | |

**Supplemental Table 7.** *Association of age and DunedinPACE controlling for white blood cell count proportions*

|  | **Bivariate association between   age and DunedinPACE** | | | **Association when  controlling for WBC** | |
| --- | --- | --- | --- | --- | --- |
| **Cohort** | **Obs.** | ***B*** | **95% CI** | ***B*** | **95% CI** |
| HRS | 3,483 | 0.0015 | 0.0010, 0.0020 | 0.0004 | -0.0001, 0.0088 |
| CARDIA | 6,214 | 0.0017 | 0.0013, 0.0021 | 0.0010 | 0.0006, 0.0014 |
| WHI | 1,845 | -0.0013 | -0.0021, -0.0005 | -0.0011 | -0.0019, -0.0004 |
| FHS Offspring | 2,673 | 0.0042 | 0.0037, 0.0048 | 0.0024 | 0.0018, 0.0029 |
| FHS 3rd Gen. | 492 | 0.0020 | 0.0010, 0.0030 | 0.0002 | -0.0009, 0.0013 |
| DNHS | 846 | 0.0019 | 0.0012, 0.0025 | 0.0005 | -0.0001, 0.0011 |
| SATSA | 1,072 | 0.0042 | 0.0030, 0.0054 | 0.0022 | 0.0010, 0.0034 |
| Add Health | 4,627 | 0.0058 | 0.0038, 0.0078 | 0.0051 | 0.0032, 0.0070 |
| PDMH | 2,309 | 0.0026 | 0.0022, 0.0031 | 0.0028 | 0.0024, 0.0032 |
| Gen. Scotland | 18,140 | 0.0024 | 0.0023, 0.0025 | 0.0026 | 0.0024, 0.0027 |
| **Meta-analyzed results** | | 0.0024 | 0.0014, 0.0034 | 0.0015 | 0.0005, 0.0025 |
| *Note*: Meta-analyzed results were generated using a random effects model. MIDUS data did not include WBCs for analysis and were not included as a result Gen. = Generation; Obs. = observations, WBCs = White blood cells. | | | | | |

**Supplemental Table 8.** DunedinPACE normed scores from age 20 to 90, overall and by sex.

|  | **Overall score** | | | | | | **For men** | | | | | **For women** | | | | |
| --- | --- | --- | --- | --- | --- | --- | --- | --- | --- | --- | --- | --- | --- | --- | --- | --- |
| **Age** | **10th%** | **30th%** | **50th%** | **70th%** | **90th%** | **10th%** | | **30th%** | **50th%** | **70th%** | **90th%** | **10th%** | **30th%** | **50th%** | **70th%** | **90th%** |
| **20** | 0.774 | 0.872 | 0.940 | 1.008 | 1.106 | 0.749 | | 0.847 | 0.915 | 0.983 | 1.081 | 0.791 | 0.889 | 0.958 | 1.026 | 1.124 |
| **21** | 0.776 | 0.874 | 0.942 | 1.011 | 1.109 | 0.752 | | 0.850 | 0.918 | 0.987 | 1.085 | 0.793 | 0.891 | 0.959 | 1.027 | 1.126 |
| **22** | 0.778 | 0.877 | 0.945 | 1.013 | 1.111 | 0.755 | | 0.854 | 0.922 | 0.990 | 1.088 | 0.795 | 0.893 | 0.961 | 1.029 | 1.127 |
| **23** | 0.781 | 0.879 | 0.947 | 1.015 | 1.114 | 0.759 | | 0.857 | 0.925 | 0.993 | 1.092 | 0.796 | 0.894 | 0.963 | 1.031 | 1.129 |
| **24** | 0.783 | 0.881 | 0.950 | 1.018 | 1.116 | 0.762 | | 0.860 | 0.929 | 0.997 | 1.095 | 0.798 | 0.896 | 0.964 | 1.032 | 1.131 |
| **25** | 0.786 | 0.884 | 0.952 | 1.020 | 1.118 | 0.766 | | 0.864 | 0.932 | 1.000 | 1.098 | 0.800 | 0.898 | 0.966 | 1.034 | 1.132 |
| **26** | 0.788 | 0.886 | 0.954 | 1.023 | 1.121 | 0.769 | | 0.867 | 0.935 | 1.004 | 1.102 | 0.801 | 0.900 | 0.968 | 1.036 | 1.134 |
| **27** | 0.790 | 0.889 | 0.957 | 1.025 | 1.123 | 0.772 | | 0.871 | 0.939 | 1.007 | 1.105 | 0.803 | 0.901 | 0.969 | 1.038 | 1.136 |
| **28** | 0.793 | 0.891 | 0.959 | 1.027 | 1.126 | 0.776 | | 0.874 | 0.942 | 1.010 | 1.109 | 0.805 | 0.903 | 0.971 | 1.039 | 1.138 |
| **29** | 0.795 | 0.893 | 0.962 | 1.030 | 1.128 | 0.779 | | 0.877 | 0.946 | 1.014 | 1.112 | 0.806 | 0.905 | 0.973 | 1.041 | 1.139 |
| **30** | 0.798 | 0.896 | 0.964 | 1.032 | 1.130 | 0.783 | | 0.881 | 0.949 | 1.017 | 1.115 | 0.808 | 0.906 | 0.975 | 1.043 | 1.141 |
| **31** | 0.800 | 0.898 | 0.966 | 1.035 | 1.133 | 0.786 | | 0.884 | 0.952 | 1.021 | 1.119 | 0.810 | 0.908 | 0.976 | 1.044 | 1.143 |
| **32** | 0.802 | 0.901 | 0.969 | 1.037 | 1.135 | 0.789 | | 0.888 | 0.956 | 1.024 | 1.122 | 0.812 | 0.910 | 0.978 | 1.046 | 1.144 |
| **33** | 0.805 | 0.903 | 0.971 | 1.039 | 1.138 | 0.793 | | 0.891 | 0.959 | 1.027 | 1.126 | 0.813 | 0.911 | 0.980 | 1.048 | 1.146 |
| **34** | 0.807 | 0.905 | 0.974 | 1.042 | 1.140 | 0.796 | | 0.894 | 0.963 | 1.031 | 1.129 | 0.815 | 0.913 | 0.981 | 1.049 | 1.148 |
| **35** | 0.810 | 0.908 | 0.976 | 1.044 | 1.142 | 0.800 | | 0.898 | 0.966 | 1.034 | 1.132 | 0.817 | 0.915 | 0.983 | 1.051 | 1.149 |
| **36** | 0.812 | 0.910 | 0.978 | 1.047 | 1.145 | 0.803 | | 0.901 | 0.969 | 1.038 | 1.136 | 0.818 | 0.917 | 0.985 | 1.053 | 1.151 |
| **37** | 0.814 | 0.913 | 0.981 | 1.049 | 1.147 | 0.806 | | 0.905 | 0.973 | 1.041 | 1.139 | 0.820 | 0.918 | 0.986 | 1.055 | 1.153 |
| **38** | 0.817 | 0.915 | 0.983 | 1.051 | 1.150 | 0.810 | | 0.908 | 0.976 | 1.044 | 1.143 | 0.822 | 0.920 | 0.988 | 1.056 | 1.155 |
| **39** | 0.819 | 0.917 | 0.986 | 1.054 | 1.152 | 0.813 | | 0.911 | 0.980 | 1.048 | 1.146 | 0.823 | 0.922 | 0.990 | 1.058 | 1.156 |
| **40** | 0.822 | 0.920 | 0.988 | 1.056 | 1.154 | 0.817 | | 0.915 | 0.983 | 1.051 | 1.149 | 0.825 | 0.923 | 0.992 | 1.060 | 1.158 |
| **41** | 0.824 | 0.922 | 0.990 | 1.059 | 1.157 | 0.820 | | 0.918 | 0.986 | 1.055 | 1.153 | 0.827 | 0.925 | 0.993 | 1.061 | 1.160 |
| **42** | 0.826 | 0.925 | 0.993 | 1.061 | 1.159 | 0.823 | | 0.922 | 0.990 | 1.058 | 1.156 | 0.829 | 0.927 | 0.995 | 1.063 | 1.161 |
| **43** | 0.829 | 0.927 | 0.995 | 1.063 | 1.162 | 0.827 | | 0.925 | 0.993 | 1.061 | 1.160 | 0.830 | 0.928 | 0.997 | 1.065 | 1.163 |
| **44** | 0.831 | 0.929 | 0.998 | 1.066 | 1.164 | 0.830 | | 0.928 | 0.997 | 1.065 | 1.163 | 0.832 | 0.930 | 0.998 | 1.066 | 1.165 |
| **45** | 0.834 | 0.932 | 1.000 | 1.068 | 1.166 | 0.834 | | 0.932 | 1.000 | 1.068 | 1.166 | 0.834 | 0.932 | 1.000 | 1.068 | 1.166 |
| **46** | 0.836 | 0.934 | 1.002 | 1.071 | 1.169 | 0.837 | | 0.935 | 1.003 | 1.072 | 1.170 | 0.835 | 0.934 | 1.002 | 1.070 | 1.168 |
| **47** | 0.838 | 0.937 | 1.005 | 1.073 | 1.171 | 0.840 | | 0.939 | 1.007 | 1.075 | 1.173 | 0.837 | 0.935 | 1.003 | 1.072 | 1.170 |
| **48** | 0.841 | 0.939 | 1.007 | 1.075 | 1.174 | 0.844 | | 0.942 | 1.010 | 1.078 | 1.177 | 0.839 | 0.937 | 1.005 | 1.073 | 1.172 |
| **49** | 0.843 | 0.941 | 1.010 | 1.078 | 1.176 | 0.847 | | 0.945 | 1.014 | 1.082 | 1.180 | 0.840 | 0.939 | 1.007 | 1.075 | 1.173 |
| **50** | 0.846 | 0.944 | 1.012 | 1.080 | 1.178 | 0.851 | | 0.949 | 1.017 | 1.085 | 1.183 | 0.842 | 0.940 | 1.009 | 1.077 | 1.175 |
| **51** | 0.848 | 0.946 | 1.014 | 1.083 | 1.181 | 0.854 | | 0.952 | 1.020 | 1.089 | 1.187 | 0.844 | 0.942 | 1.010 | 1.078 | 1.177 |
| **52** | 0.850 | 0.949 | 1.017 | 1.085 | 1.183 | 0.857 | | 0.956 | 1.024 | 1.092 | 1.190 | 0.846 | 0.944 | 1.012 | 1.080 | 1.178 |
| **53** | 0.853 | 0.951 | 1.019 | 1.087 | 1.186 | 0.861 | | 0.959 | 1.027 | 1.095 | 1.194 | 0.847 | 0.945 | 1.014 | 1.082 | 1.180 |
| **54** | 0.855 | 0.953 | 1.022 | 1.090 | 1.188 | 0.864 | | 0.962 | 1.031 | 1.099 | 1.197 | 0.849 | 0.947 | 1.015 | 1.083 | 1.182 |
| **55** | 0.858 | 0.956 | 1.024 | 1.092 | 1.190 | 0.868 | | 0.966 | 1.034 | 1.102 | 1.200 | 0.851 | 0.949 | 1.017 | 1.085 | 1.183 |
| **56** | 0.860 | 0.958 | 1.026 | 1.095 | 1.193 | 0.871 | | 0.969 | 1.037 | 1.106 | 1.204 | 0.852 | 0.951 | 1.019 | 1.087 | 1.185 |
| **57** | 0.862 | 0.961 | 1.029 | 1.097 | 1.195 | 0.874 | | 0.973 | 1.041 | 1.109 | 1.207 | 0.854 | 0.952 | 1.020 | 1.089 | 1.187 |
| **58** | 0.865 | 0.963 | 1.031 | 1.099 | 1.198 | 0.878 | | 0.976 | 1.044 | 1.112 | 1.211 | 0.856 | 0.954 | 1.022 | 1.090 | 1.189 |
| **59** | 0.867 | 0.965 | 1.034 | 1.102 | 1.200 | 0.881 | | 0.979 | 1.048 | 1.116 | 1.214 | 0.857 | 0.956 | 1.024 | 1.092 | 1.190 |
| **60** | 0.870 | 0.968 | 1.036 | 1.104 | 1.202 | 0.885 | | 0.983 | 1.051 | 1.119 | 1.217 | 0.859 | 0.957 | 1.026 | 1.094 | 1.192 |
| **61** | 0.872 | 0.970 | 1.038 | 1.107 | 1.205 | 0.888 | | 0.986 | 1.054 | 1.123 | 1.221 | 0.861 | 0.959 | 1.027 | 1.095 | 1.194 |
| **62** | 0.874 | 0.973 | 1.041 | 1.109 | 1.207 | 0.891 | | 0.990 | 1.058 | 1.126 | 1.224 | 0.863 | 0.961 | 1.029 | 1.097 | 1.195 |
| **63** | 0.877 | 0.975 | 1.043 | 1.111 | 1.210 | 0.895 | | 0.993 | 1.061 | 1.129 | 1.228 | 0.864 | 0.962 | 1.031 | 1.099 | 1.197 |
| **64** | 0.879 | 0.977 | 1.046 | 1.114 | 1.212 | 0.898 | | 0.996 | 1.065 | 1.133 | 1.231 | 0.866 | 0.964 | 1.032 | 1.100 | 1.199 |
| **65** | 0.882 | 0.980 | 1.048 | 1.116 | 1.214 | 0.902 | | 1.000 | 1.068 | 1.136 | 1.234 | 0.868 | 0.966 | 1.034 | 1.102 | 1.200 |
| **66** | 0.884 | 0.982 | 1.050 | 1.119 | 1.217 | 0.905 | | 1.003 | 1.071 | 1.140 | 1.238 | 0.869 | 0.968 | 1.036 | 1.104 | 1.202 |
| **67** | 0.886 | 0.985 | 1.053 | 1.121 | 1.219 | 0.908 | | 1.007 | 1.075 | 1.143 | 1.241 | 0.871 | 0.969 | 1.037 | 1.106 | 1.204 |
| **68** | 0.889 | 0.987 | 1.055 | 1.123 | 1.222 | 0.912 | | 1.010 | 1.078 | 1.146 | 1.245 | 0.873 | 0.971 | 1.039 | 1.107 | 1.206 |
| **69** | 0.891 | 0.989 | 1.058 | 1.126 | 1.224 | 0.915 | | 1.013 | 1.082 | 1.150 | 1.248 | 0.874 | 0.973 | 1.041 | 1.109 | 1.207 |
| **70** | 0.894 | 0.992 | 1.060 | 1.128 | 1.226 | 0.919 | | 1.017 | 1.085 | 1.153 | 1.251 | 0.876 | 0.974 | 1.043 | 1.111 | 1.209 |
| **71** | 0.896 | 0.994 | 1.062 | 1.131 | 1.229 | 0.922 | | 1.020 | 1.088 | 1.157 | 1.255 | 0.878 | 0.976 | 1.044 | 1.112 | 1.211 |
| **72** | 0.898 | 0.997 | 1.065 | 1.133 | 1.231 | 0.925 | | 1.024 | 1.092 | 1.160 | 1.258 | 0.880 | 0.978 | 1.046 | 1.114 | 1.212 |
| **73** | 0.901 | 0.999 | 1.067 | 1.135 | 1.234 | 0.929 | | 1.027 | 1.095 | 1.163 | 1.262 | 0.881 | 0.979 | 1.048 | 1.116 | 1.214 |
| **74** | 0.903 | 1.001 | 1.070 | 1.138 | 1.236 | 0.932 | | 1.030 | 1.099 | 1.167 | 1.265 | 0.883 | 0.981 | 1.049 | 1.117 | 1.216 |
| **75** | 0.906 | 1.004 | 1.072 | 1.140 | 1.238 | 0.936 | | 1.034 | 1.102 | 1.170 | 1.268 | 0.885 | 0.983 | 1.051 | 1.119 | 1.217 |
| **76** | 0.908 | 1.006 | 1.074 | 1.143 | 1.241 | 0.939 | | 1.037 | 1.105 | 1.174 | 1.272 | 0.886 | 0.985 | 1.053 | 1.121 | 1.219 |
| **77** | 0.910 | 1.009 | 1.077 | 1.145 | 1.243 | 0.942 | | 1.041 | 1.109 | 1.177 | 1.275 | 0.888 | 0.986 | 1.054 | 1.123 | 1.221 |
| **78** | 0.913 | 1.011 | 1.079 | 1.147 | 1.246 | 0.946 | | 1.044 | 1.112 | 1.180 | 1.279 | 0.890 | 0.988 | 1.056 | 1.124 | 1.223 |
| **79** | 0.915 | 1.013 | 1.082 | 1.150 | 1.248 | 0.949 | | 1.047 | 1.116 | 1.184 | 1.282 | 0.891 | 0.990 | 1.058 | 1.126 | 1.224 |
| **80** | 0.918 | 1.016 | 1.084 | 1.152 | 1.250 | 0.953 | | 1.051 | 1.119 | 1.187 | 1.285 | 0.893 | 0.991 | 1.060 | 1.128 | 1.226 |
| **81** | 0.920 | 1.018 | 1.086 | 1.155 | 1.253 | 0.956 | | 1.054 | 1.122 | 1.191 | 1.289 | 0.895 | 0.993 | 1.061 | 1.129 | 1.228 |
| **82** | 0.922 | 1.021 | 1.089 | 1.157 | 1.255 | 0.959 | | 1.058 | 1.126 | 1.194 | 1.292 | 0.897 | 0.995 | 1.063 | 1.131 | 1.229 |
| **83** | 0.925 | 1.023 | 1.091 | 1.159 | 1.258 | 0.963 | | 1.061 | 1.129 | 1.197 | 1.296 | 0.898 | 0.996 | 1.065 | 1.133 | 1.231 |
| **84** | 0.927 | 1.025 | 1.094 | 1.162 | 1.260 | 0.966 | | 1.064 | 1.133 | 1.201 | 1.299 | 0.900 | 0.998 | 1.066 | 1.134 | 1.233 |
| **85** | 0.930 | 1.028 | 1.096 | 1.164 | 1.262 | 0.970 | | 1.068 | 1.136 | 1.204 | 1.302 | 0.902 | 1.000 | 1.068 | 1.136 | 1.234 |
| **86** | 0.932 | 1.030 | 1.098 | 1.167 | 1.265 | 0.973 | | 1.071 | 1.139 | 1.208 | 1.306 | 0.903 | 1.002 | 1.070 | 1.138 | 1.236 |
| **87** | 0.934 | 1.033 | 1.101 | 1.169 | 1.267 | 0.976 | | 1.075 | 1.143 | 1.211 | 1.309 | 0.905 | 1.003 | 1.071 | 1.140 | 1.238 |
| **88** | 0.937 | 1.035 | 1.103 | 1.171 | 1.270 | 0.980 | | 1.078 | 1.146 | 1.214 | 1.313 | 0.907 | 1.005 | 1.073 | 1.141 | 1.240 |
| **89** | 0.939 | 1.037 | 1.106 | 1.174 | 1.272 | 0.983 | | 1.081 | 1.150 | 1.218 | 1.316 | 0.908 | 1.007 | 1.075 | 1.143 | 1.241 |
| **90** | 0.942 | 1.040 | 1.108 | 1.176 | 1.274 | 0.987 | | 1.085 | 1.153 | 1.221 | 1.319 | 0.910 | 1.008 | 1.077 | 1.145 | 1.243 |

**Supplemental Text 2**

**Age-Normed Reference Scores.** Combining the sex-specific linear age parameters generated in our results with the DunedinPACE reference score for 45-year-olds (1.00) allows for a simple calculation of age-normed reference scores:

Men: *Age-Normed Reference Score* = [(Age – 45) * .0034] + 1.00)

Women: *Age-Normed Reference Score* = [(Age – 45) * .0017] + 1.00)

These scores represent the predicted average aging expected for any age across the adult lifespan.

**Variation in DunedinPACE.** In addition to reference values, norms also require expected variation for those scores. The average variation in DunedinPACE was *SD* = 0.13 across cohorts, with remarkably similar variation for men (*SD* = 0.13) and women (*SD* = 0.13) and across age periods (**Supplemental Table 9)**. As a result, we used a *SD* of 0.13 for the norms across the adult lifespan. Combining the normed scores and variation in DunedinPACE allows for a calculation of *z*-scores (and associated percentile ranks) for DunedinPACE as follows:

*Pace of Aging Z-Score* = (DunedinPACE - *Age-Normed Reference Score*) / 0.13

Age-normed scores for men and women are illustrated in **Figure 2**, whereas **Supplemental Figure 1** illustrates these scores for combined samples that do not (or cannot) account for sex-specific differences. Values with different percentile ranks in **Supplemental Table 8**.

**Generating 95% Confidence Intervals for DunedinPACE Aging Scores.** We combined the weighted average *SD* with the test-retest reliability of DunedinPACE from the original publication in 2022 (ICC = 0.965) to calculate the standard error of the measurement (SE_m_) for any given DunedinPACE aging score (SE_m_ = 0.0243). Rounding this to a more conservative 0.025 and doubling the value produced a 95% confidence interval around any DunedinPACE aging score (± 0.05). This confidence interval allows someone receiving a DunedinPACE score of 1.06 to be told that if their DunedinPACE was measured again, 95 out of 100 times their value would lie between 1.01 and 1.11

**Supplemental Table 9.** *Variance in DunedinPACE across cohorts, men and women, and age ranges*

|  | **Standard deviations (*SD*s)** | | | | | | | |
| --- | --- | --- | --- | --- | --- | --- | --- | --- |
| **Cohort** | **Overall** | **Men** | **Women** | **Ages 18-44** | **Age 45-64** | | | **Age 65+** |
| HRS | 0.14 | 0.13 | 0.14 | — | | 0.14 | 0.13 | |
| CARDIA | 0.13 | 0.13 | 0.14 | 0.13 | | 0.13 | — | |
| WHI | 0.12 | — | 0.12 | — | | 0.13 | 0.12 | |
| FHS Offspring | 0.13 | 0.13 | 0.13 | — | | 0.12 | 0.12 | |
| FHS 3rd Gen. | 0.11 | 0.11 | 0.11 | — | | 0.11 | — | |
| DNHS | 0.13 | 0.13 | 0.13 | — | | 0.13 | 0.13 | |
| SATSA | 0.17 | 0.17 | 0.17 | — | | 0.15 | 0.17 | |
| Add Health | 0.13 | 0.13 | 0.13 | 0.13 | | — | — | |
| MIDUS | 0.14 | 0.14 | 0.14 | 0.14 | | 0.14 | 0.13 | |
| PDMH | 0.11 | 0.11 | 0.11 | 0.11 | | 0.11 | — | |
| Generation Scotland | 0.13 | 0.13 | 0.13 | 0.12 | | 0.13 | 0.13 | |
| **Weighted averages** | **0.130** | **0.129** | **0.132** | **0.123** | | **0.130** | **0.131** | |
| *Note*: *SD* totals = weighted averages based on sample size for the cohorts. Age ranges were selected *a priori* to match with years of life associated with adulthood, midlife, and older age broadly defined. | | | | | | | | |

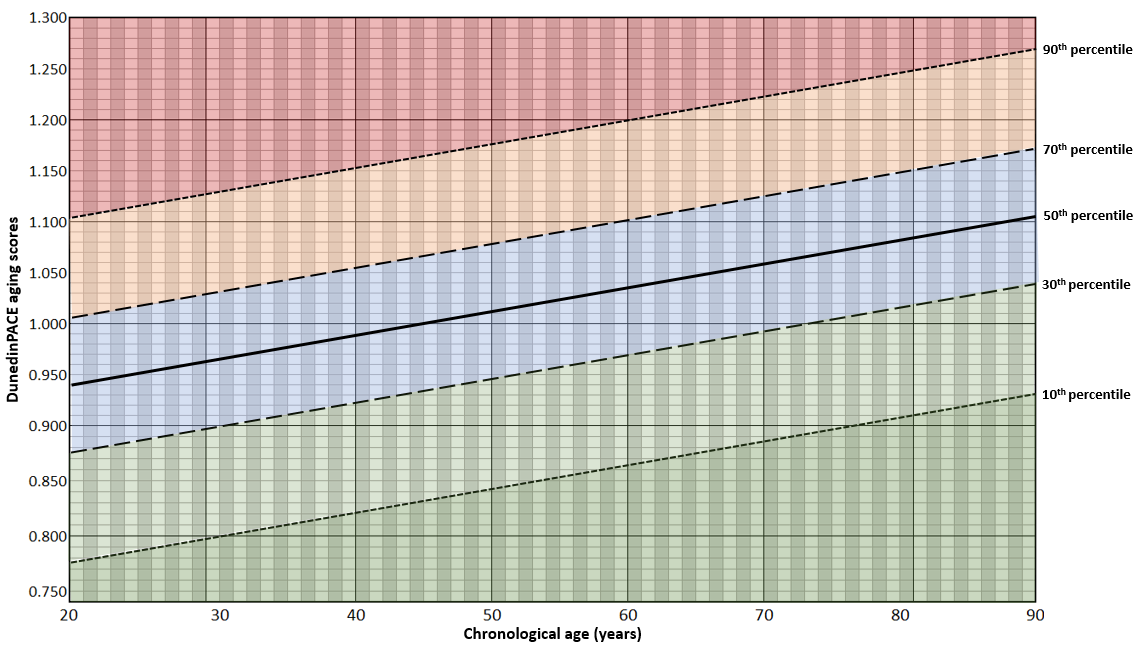

**Supplemental Figure 1.** Illustration of the DunedinPACE age norms and percentiles around the normed values at each age from 20 to 90 years old. Values were calculated using the same approach as for the sex-specific norms (Age-normed DunedinPACE = [(Age – 45) * .0024] + 1.00) with the exception that the overall meta-analysis value is used to create the reference line.

**Supplemental Table 10.** *Comparing the association with future disease and death for the original DunedinPACE and age-normed DunedinPACE*

|  | 10-year change in  chronic disease | | | Onset of any  chronic disease | | | All-cause  mortality | | |
| --- | --- | --- | --- | --- | --- | --- | --- | --- | --- |
|  | **β** | **95% CI** | ***p*** | ***HR*** | **95% CI** | ***p*** | ***HR*** | **95% CI** | ***p*** |
| **Models run in PDMH** |  |  |  |  |  |  |  |  |  |
| Original DunedinPACE | 0.33 | 0.26-0.40 | < .001 | 1.41 | 1.31-1.52 | < .001 | 1.36 | 1.13-1.64 | .001 |
| Age-normed DunedinPACE | 0.36 | 0.29-0.44 | < .001 | 1.45 | 1.33-1.57 | < .001 | 1.38 | 1.13-1.69 | .002 |
| **Models run in MIDUS** |  |  |  |  |  |  |  |  |  |
| Original DunedinPACE |  |  |  |  |  |  | 1.96 | 1.67-2.30 | < .001 |
| Age-normed DunedinPACE |  |  |  |  |  |  | 2.02 | 1.71-2.40 | < .001 |
| *Note*: All models included chronological age as a covariate. PDMH models included 2,265 veterans with EHR data and the MIDUS study models included 1,309 individuals. All models used either negative binomial regression (for change in chronic disease burden, to account for the distribution of chronic disease counts) or Cox-proportional hazard models predicting time to event (for onset of chronic disease and all-cause mortality). The PDMH models included 630 individuals who developed a chronic disease and 102 individuals who died over follow-up. The MIDUS study included 150 individuals who died over follow-up. Models predicting 10-year chronic disease change excluded 182 participants who did not have 10 years of health record observation, models predicting chronic disease onset in the PDMH excluded 301 individuals with a chronic disease already diagnoses at baseline. Age-normed DunedinPACE scores were created using the sex-specific age-normed DunedinPACE calculations. Models for 10-year change in chronic disease included baseline chronic disease burden as a predictor. Outcomes are scaled to 1 *SD* unit in DunedinPACE. | | | | | | | | | |

**Supplemental Methods 2. Detailed description of clinical outcomes derived in the PDMH and MIDUS**

Prospective health outcomes and clinical biomarkers were derived using data from Veteran’s Affairs electronic health records. Veterans enrolled in the Post-Deployment Mental Health^1^ (PDMH) study with a baseline that ranged from 2005 to 2016, which resulted in follow-up periods that ranged from 8.4 to 19.5 years. Data linkage was completed in the VA Informatics and Computing Infrastructure (VINCI) system using social security numbers (SSNs) from the PDMH linked to VA patient’s social security and internal control numbers (ICN). Data for the relevant health outcomes were then called from VA Corporate Data Warehouse (CDW) tables using SQL coding. Chronic disease and biomarker data were generated from outpatient, inpatient, and purchased care data (e.g., community care that is delivered to veterans following referral from the VA and/or paid by VA sources). Data cleaning processes are reported in the following sections**.** Data pulls were completed on 9/26/2025 to a censor date of 12/31/2024, resulting an average of 14.1 years of assessment.

***Charlson Comorbidity Index Scores.*** Charlson comorbidity index (CCI) scores were derived using diagnostic ICD-9 and ICD-10 codes^2^, matching the tables crated by Glasheen and colleagues. Baseline values used ICD codes at the date of enrollment in the PDMH and were updated for each subsequent year. Chronic diseases diagnoses were carried forward to subsequent years. Scores were coded as missing for any year past the end of the veteran’s follow-up observation period, as well as if there was insufficient evidence of a patient utilizing VA healthcare and no diagnoses were present. Presence of chronic diseases were defined as a diagnosis being included in two outpatient or one outpatient visit. Chronic disease diagnostic statuses for the major categories of chronic disease included in the CCI were ascertained by presence of ICD codes in veterans’ EHR drawn directly from tables outlined by Glasheen and colleagues^2^. The onset of any major chronic disease represented the onset of any chronic disease.

**Mortality.** All-cause mortality was ascertained using data from the VA CDW’s death ascertainment file to the end of 2023. In MIDUS, mortality was ascertained in the MIDUS Core and Refresher Mortality datasets^30,31^ to the end of 2023. The date of biomarker data collection was combined with the date of death or censor date to produce the measure of time.

**Supplemental References 4**

1. Brancu M, Wagner HR, Morey RA, et al. The Post-Deployment Mental Health (PDMH) study and repository: A multi-site study of US Afghanistan and Iraq era veterans. *Int J Methods Psychiatr Res*. 2017;26(3):e1570. doi:10.1002/mpr.1570
2. Glasheen WP, Cordier T, Gumpina R, Haugh G, Davis J, Renda A. Charlson Comorbidity Index: *ICD-9* Update and *ICD-10* Translation. *Am Health Drug Benefits*. 2019;12(4):188-197.
3. Ryff, Carol D., Almeida, David, Ayanian, John Z., Binkley, Neil, Carr, Deborah S., Coe, Christopher, … Williams, David R. Midlife in the United States: Core Sample Mortality Data, 1995-2023. Inter-university Consortium for Political and Social Research [distributor], 2025-07-10. https://doi.org/10.3886/ICPSR37237.v6
4. Ryff, Carol D., Almeida, David M., Ayanian, John Z., Binkley, Neil, Carr, Deborah S., Coe, Christopher, …Williams, David R. Midlife in the United States: Refresher Sample Mortality Data, 2012-2023. Inter-university Consortium for Political and Social Research [distributor], 2025-05-28. https://doi.org/10.3886/ICPSR38024.v3
